# Development and Validation of Multivariable Prediction Models of Serological Response to SARS-CoV-2 Vaccination in Kidney Transplant Recipients

**DOI:** 10.1101/2022.06.02.22275894

**Authors:** Bilgin Osmanodja, Johannes Stegbauer, Marta Kantauskaite, Lars Christian Rump, Andreas Heinzel, Roman Reindl-Schwaighofer, Rainer Oberbauer, Ilies Benotmane, Sophie Caillard, Christophe Masset, Clarisse Kerleau, Gilles Blancho, Klemens Budde, Fritz Grunow, Michael Mikhailov, Eva Schrezenmeier, Simon Ronicke

## Abstract

**Background:** Repeated vaccination against SARS-CoV-2 increases serological response in kidney transplant recipients (KTR) with high interindividual variability. No decision support tool exists to predict SARS-CoV-2 vaccination response in KTR.

**Methods:** We developed, internally and externally validated five different multivariable prediction models of serological response after the third and fourth vaccine dose against SARS-CoV-2 in KTR. Using 27 candidate predictor variables, we applied statistical and machine learning approaches including logistic regression (LR), LASSO-regularized LR, random forest, and gradient boosted regression trees. For development and internal validation, data from 585 vaccinations were used. External validation was performed in four independent, international validation datasets comprising 191, 184, 254, and 323 vaccinations, respectively.

**Findings:** LASSO-regularized LR performed on the whole development dataset yielded a 23- and 11- variable model, respectively. External validation showed AUC-ROC of 0.855, 0.749, 0.828, and 0.787 for the sparser 11-variable model, yielding an overall performance 0.819.

**Interpretation:** An 11-variable LASSO-regularized LR model predicts vaccination response in KTR with good overall accuracy. Implemented as an online tool, it can guide decisions when choosing between different immunization strategies to improve protection against COVID-19 in KTR.

## Introduction

SARS-CoV-2 vaccination offers protection from severe coronavirus disease 2019 (COVID-19) regardless of the causative variant for most healthy individuals.^1^ In contrast, protection in immunocompromised solid organ transplant (SOT) recipients is limited. The serological response rate after SARS CoV-2 vaccination in kidney transplant recipients (KTR) after three doses of vaccine is strongly impaired in comparison to the general population – resulting in insufficient protection and a COVID-19 mortality which is unacceptable high within this population.^2,3^

Different strategies to induce humoral protection for KTR have been suggested, including repeated vaccination and vaccination under adjusted immunosuppression – besides SARS-CoV-2-specific monoclonal antibody therapy.^4^ Existing data comprises risk factors identified through multivariable analyses, which are helpful to identify factors associated with insufficient vaccination response but are not easily interpretable for the single patient or vaccination.^5-7^ Specifically, no tool exists to predict response to a vaccination in a particular patient. Risk calculators can build a bridge to help assess the likelihood of vaccination success in an individual and help decide between different possible actions to take such as passive or active immunization or adjustment of immunosuppressive medication. To date, no such decision support system is available.

For this reason, we aim to develop a classification model to predict serological response to third and fourth SARS-CoV-2 vaccinations in KTR. The model’s implementation objective is to identify patients that will likely not respond to an additional dose of vaccine, even with changes in immunosuppressive medication, and thus benefit most from passive immunization strategies. Using our previously reported data of vaccination outcomes in KTR, we develop and compare a set of prediction models based on classical statistical methods as well as machine learning. After selecting the most promising models, we validate the resulting prediction models in four independent validation cohorts, with the intent to make the result available as an online risk calculator.

## Methods

### Development cohort

Data from KTR at Charité – Universitätsmedizin Berlin, Germany, were used to form the development cohort. Details of the underlying patient population, as well as the assays and cutoffs used have been previously reported.^5^ Briefly, KTR received up to five doses of SARS-CoV-2 vaccine in case of sustained lack of sufficient serological response to vaccination at our institution, usually combined with reduction or pausing MPA for fourth and fifth vaccination. For the enzyme-linked immunosorbent assays (ELISA) for the detection of IgG antibodies against the S1 domain of the SARS-CoV-2 spike (S) protein in serum (Anti-SARS-CoV-2-ELISA (IgG), EUROIMMUN Medizinische Labordiagnostika AG, Lübeck, Germany), samples with a cutoff index ≥ 1.1 (in comparison to the previously obtained cut-off value of the calibrator) were considered to be positive, samples with a cutoff index ≥ 0.8, and < 1.1 were considered low positive, and samples with a cutoff index <0.8 were considered negative, as suggested by the manufacturer.

Alternatively, for the electrochemiluminescence immunoassay (ECLIA) (Elecsys, Anti-SARS-CoV-2, Roche Diagnostics GmbH, Mannheim, Germany) detecting human immunoglobulins, including IgG, IgA and IgM against the spike receptor binding (RBD) domain protein, samples with ≥ 264 U/ml were considered to be positive as recommended by Caillard et al.^8,9^ Any non-zero antibody level below this cutoff was considered low positive (with limit of detection being 0.4 U/mL).

For predictive modeling, we included data on third and fourth vaccination, since basic immunization has most likely been performed in most KTR patients already, and since only few patients received fifth vaccination so far.

After applying all exclusion criteria summarized in **Table 1**, the development cohort included 585 vaccinations performed between December 2020 and January 2022 in 421 COVID-naive adult KTR (**Figure 1**). The Charité institutional review board approved this retrospective analysis (EA1/030/22).

**Table 1.** Inclusion and exclusion criteria regarding vaccinations.

|  |
| --- |
| **Inclusion Criteria** |
| - Functioning kidney transplant at the time of vaccination |
| - Patient 18 years or older at the time of vaccination |
| - Third or fourth SARS-CoV-2 vaccination |
| - anti-SARS-CoV-2-S-protein antibodies below positivity cutoff before respective vaccination |
| - Follow-up anti-SARS-CoV-2-S-protein antibody measurement at least 14 days after vaccination |
| **Exclusion Criteria** |
| - SARS-CoV-2 vaccinations, which were performed before transplantation or after graft loss |
| - SARS-CoV-2 infection before the vaccination or before the measurement of the respective serological response as defined by<br>- Positive SARS-CoV-2 RNA PCR<br>- Positive anti-SARS-CoV-2-N-protein antibodies |
| - Sufficient serological response before respective SARS-CoV-2 vaccination |
| - Monoclonal anti-SARS-CoV-2-S-protein antibody therapy before the measurement of the respective serological response |
| - Missing data on serological response before respective SARS-CoV-2 vaccination |
| - Missing data on serological response after respective SARS-CoV-2 vaccination |
| - Missing data on the assay used to measure serological response |
| - Missing data on immunosuppressive medication at the time of vaccination |
| - Missing lymphocyte count, eGFR, hemoglobin level |

**Figure 1.**
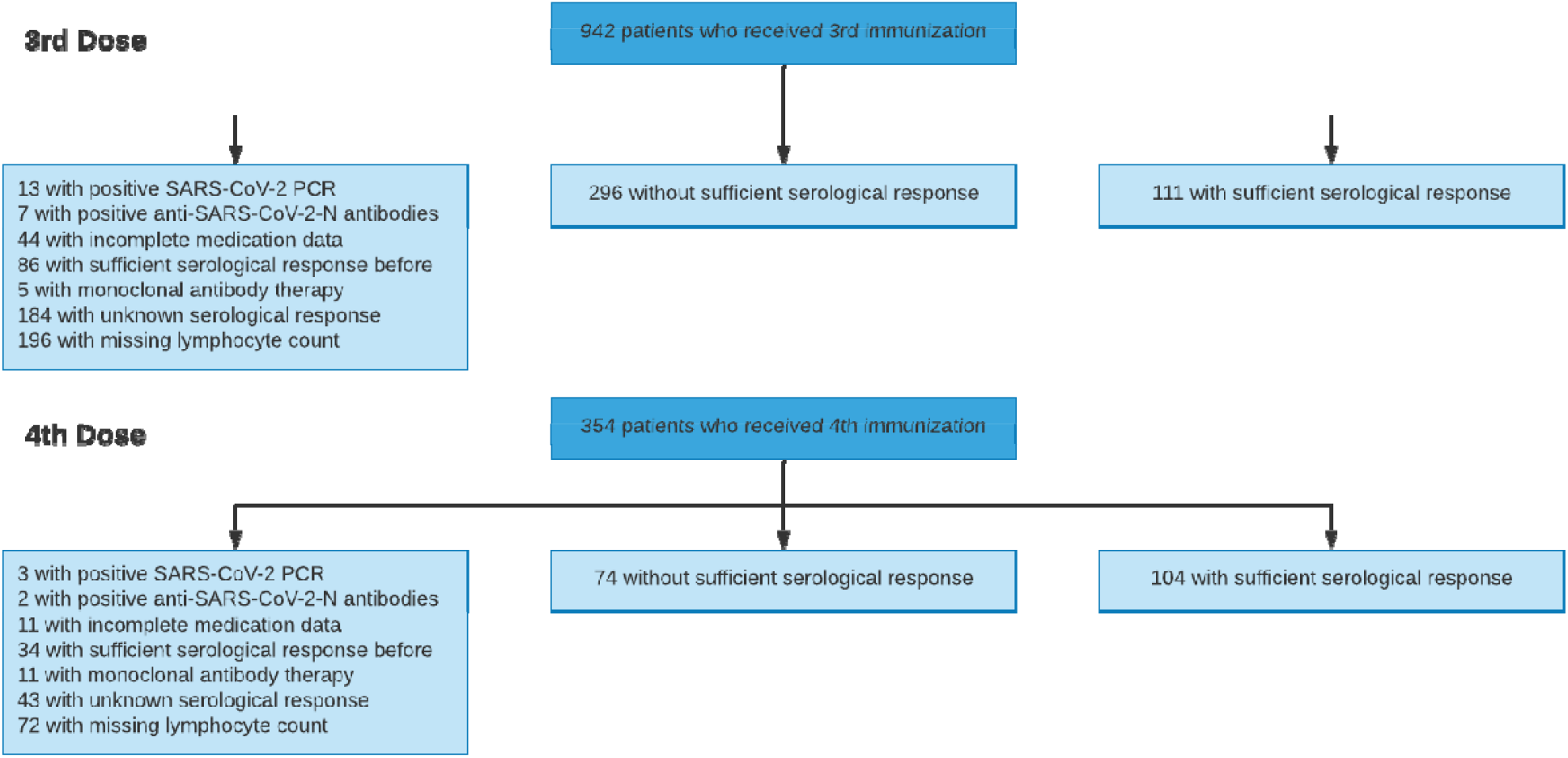
Patient flow diagram of the development cohort.

### Validation cohorts

We used four independent, international validation cohorts from outpatient transplant centers at University Hospital Düsseldorf, Germany (191 vaccinations in 137 KTR) ^10,11^, Medical University Vienna, Austria (184 vaccinations in 184 KTR) ^12^, Strasbourg University Hospital, France (254 vaccinations in 229 KTR) ^13,14^, Hotel Dieu Nantes, France (323 vaccinations in 269 KTR) ^15^. Detailed information about the validation cohorts are presented in **Items S1-S4** and patient selection including outcome frequencies are summarized separately for each validation cohort in **Figures S1-S4**.

No sample size calculation was applicable for this post-hoc analysis.

### Outcome and Predictors

The single outcome variable was a positive serological response defined by the maximum anti-SARS-CoV-2 spike (S) IgG or antibody level after a minimum of 14 days following the date of vaccination and before any further immunization event such as SARS-CoV-2 infection, passive or active immunization. Since different assays were used at different sites, details on the tests and the respective cutoffs used are provided for each validation cohort in **Item S1-S4**, which are summarized in **Table 2**. Generally, IgG or antibody positivity was determined based on local laboratory’s pre-defined positivity cutoff, which was mostly the one provided by the manufacturer. Especially for the ECLIA Elecsys assay different cutoffs were available and used, which impeded comparability. We chose to assess model performance for two cutoffs for this specific assay. First, we used the 0.8 U/mL cutoff provided by the manufacturer, yielding highest sensitivity in detecting patients with previous COVID-19. Second, a cutoff of 15 U/mL, which was initially suggested by the manufacturer to exhibit a positive predictive value of more than 99% for presence of neutralizing antibodies against the wild-type virus, was used.^12^ Contrary to the manufacturer’s designated use, our intention was to provide an alternative positivity cutoff, below which no neutralization against omicron variant occurs, but that is not as close to the limit of detection (0.4 U/mL) as the positivity cutoff provided by the manufacturer (0.8 U/mL). This alternative positivity cutoff definition was needed to test the hypothesis that indeed the absence or low number of “low-positive” antibody levels before vaccination (below the positivity cutoff, but above the limit of detection) for the ECLIA Elecsys assay led to low performance in validation sets 2 and 4. While the cutoff of 15 U/mL is somewhat arbitrary, it meets both needs mentioned above. First, it increases the percentage of low positive patients in validation set 4, and second, patients with antibody levels <50 U/mL in this assay show no neutralization against omicron BA.1, which most likely applies to omicron BA.2 as well.^16,17^ Hence, adjusting the cutoff to 15 U/mL is compatible with the objective to identify patients without serological response to an additional vaccine dose corresponding best with a lack of neutralizing antibodies.

**Table 2.**
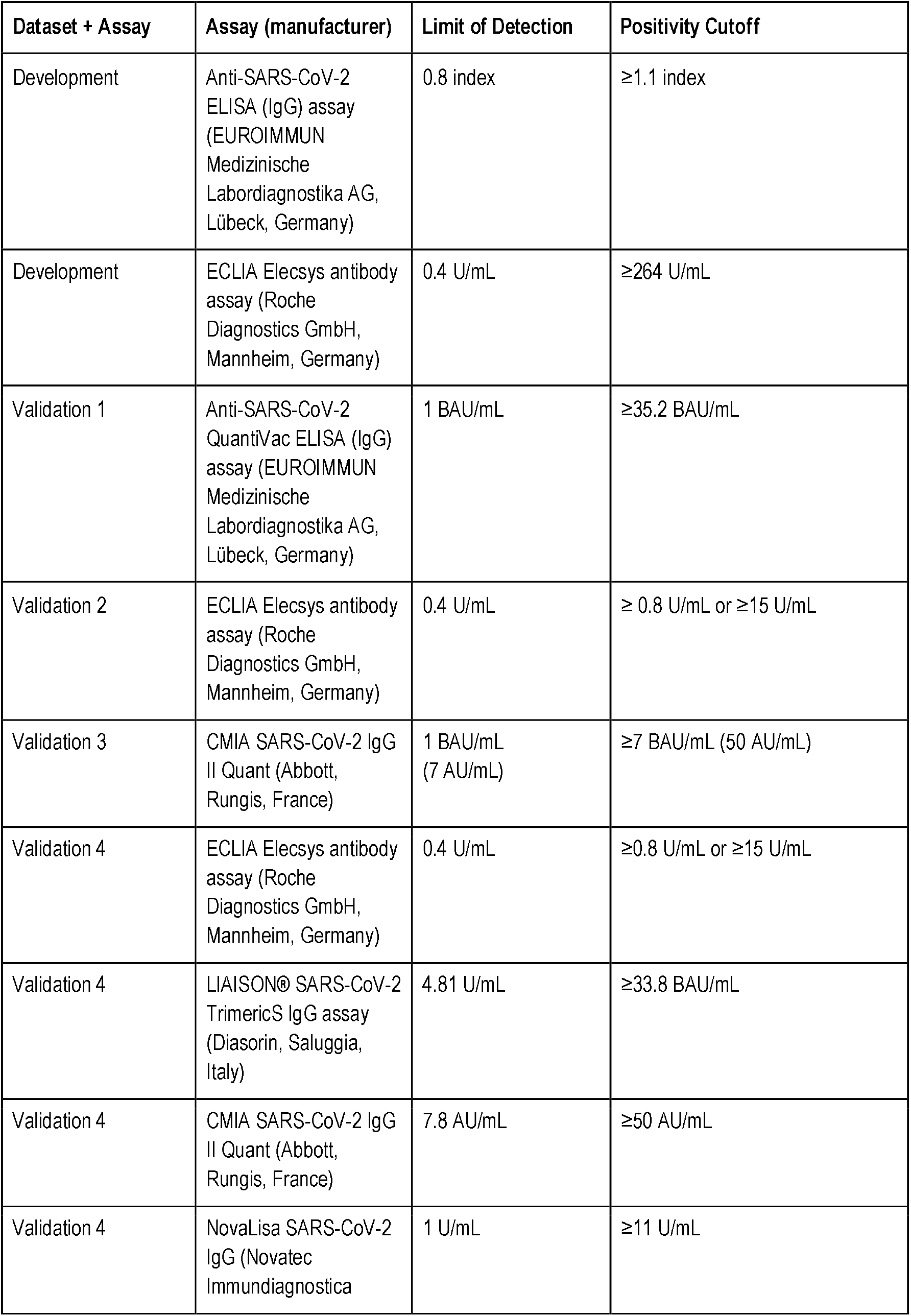
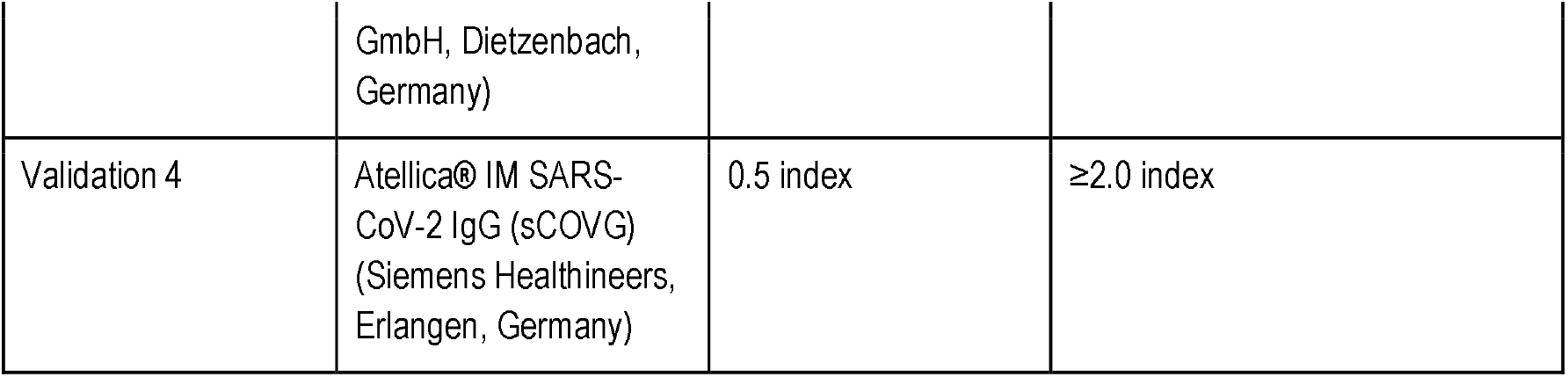
Assays, as well as respective limit of detection and positivity cutoff used for each dataset.

Predictor variables in the data sets comprised 27 variables: four vaccination-specific, three demographic, one comorbidity, four transplantation-specific, nine encoding medication, and 6 biomarkers (**Table S1**). We did not perform feature selection based on a-priori hypotheses.

### Missing data / imputation

For the development dataset, we excluded all vaccinations with incomplete data (complete case analysis). Preliminary analysis showed that neither using data from patients without lymphocyte count, which was the most common missing laboratory value, nor imputation of missing laboratory values by multiple imputation (both of which yielded higher sample size) did add predictive accuracy for logistic regression and was therefore not followed for the main analysis (**Figure S5 and Table S2**).

For each validation set, we excluded vaccinations with missing SARS-CoV-2 IgG data, missing information about the SARS-CoV-2 spike IgG or antibody assay used, missing medication data, or missing eGFR, lymphocyte count, or hemoglobin level. We chose to impute the remaining variables to reduce the number of omitted cases due to missing values. Instead of performing multiple imputation, we chose a more pragmatic approach and imputed either the most frequent value of the respective variable in the development dataset in case of binary or categorical variables, or the median (or mean) of the respective variable in the development dataset in case of numerical variables, as summarized in **Table 3**. This is the way a clinician would handle a missing value when using the online risk calculator, those values are used as presets in the online calculator. In the validation cohorts, no data originating from a time after the respective vaccination was included to make predictions.

**Table 3.**
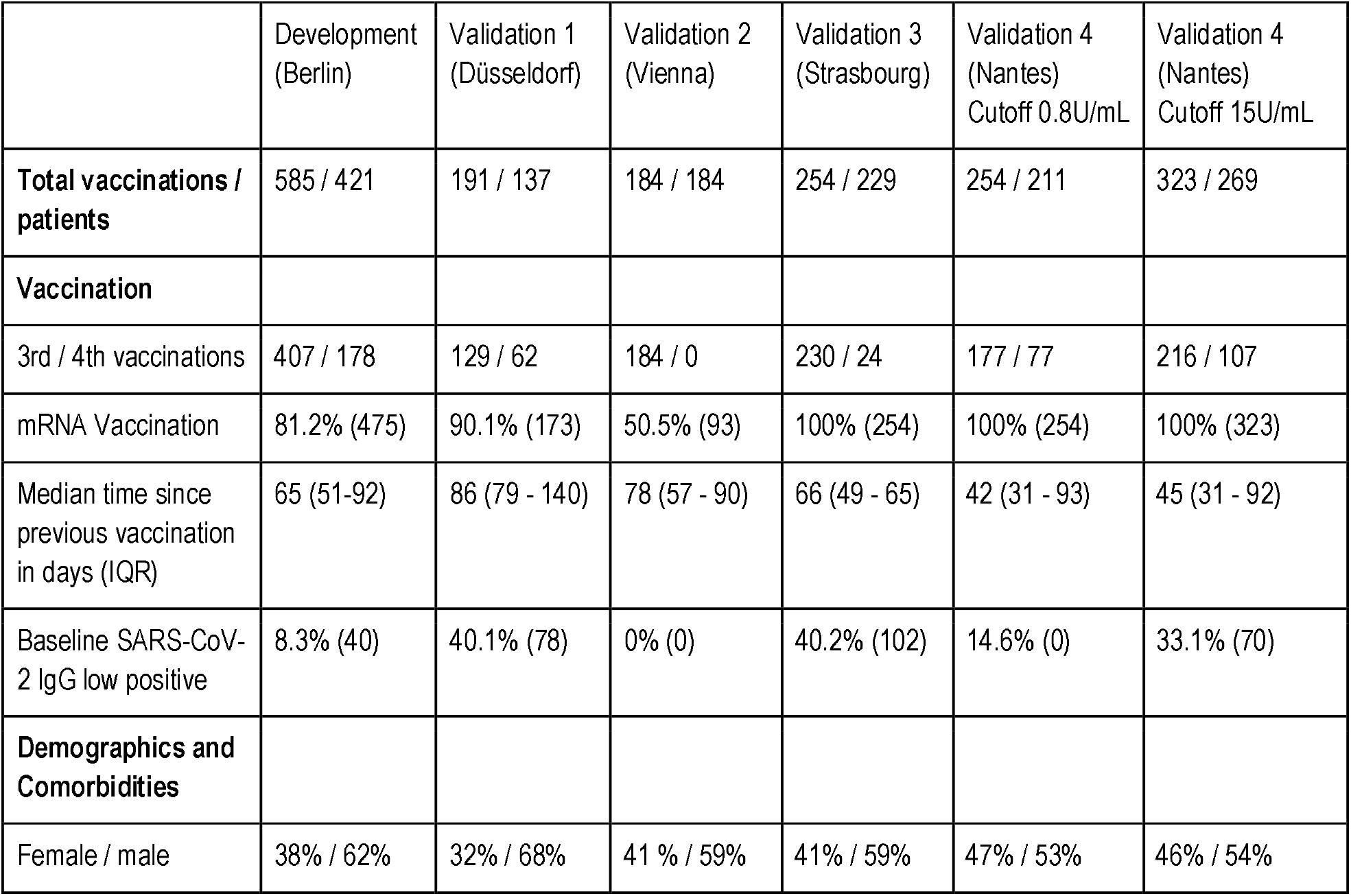
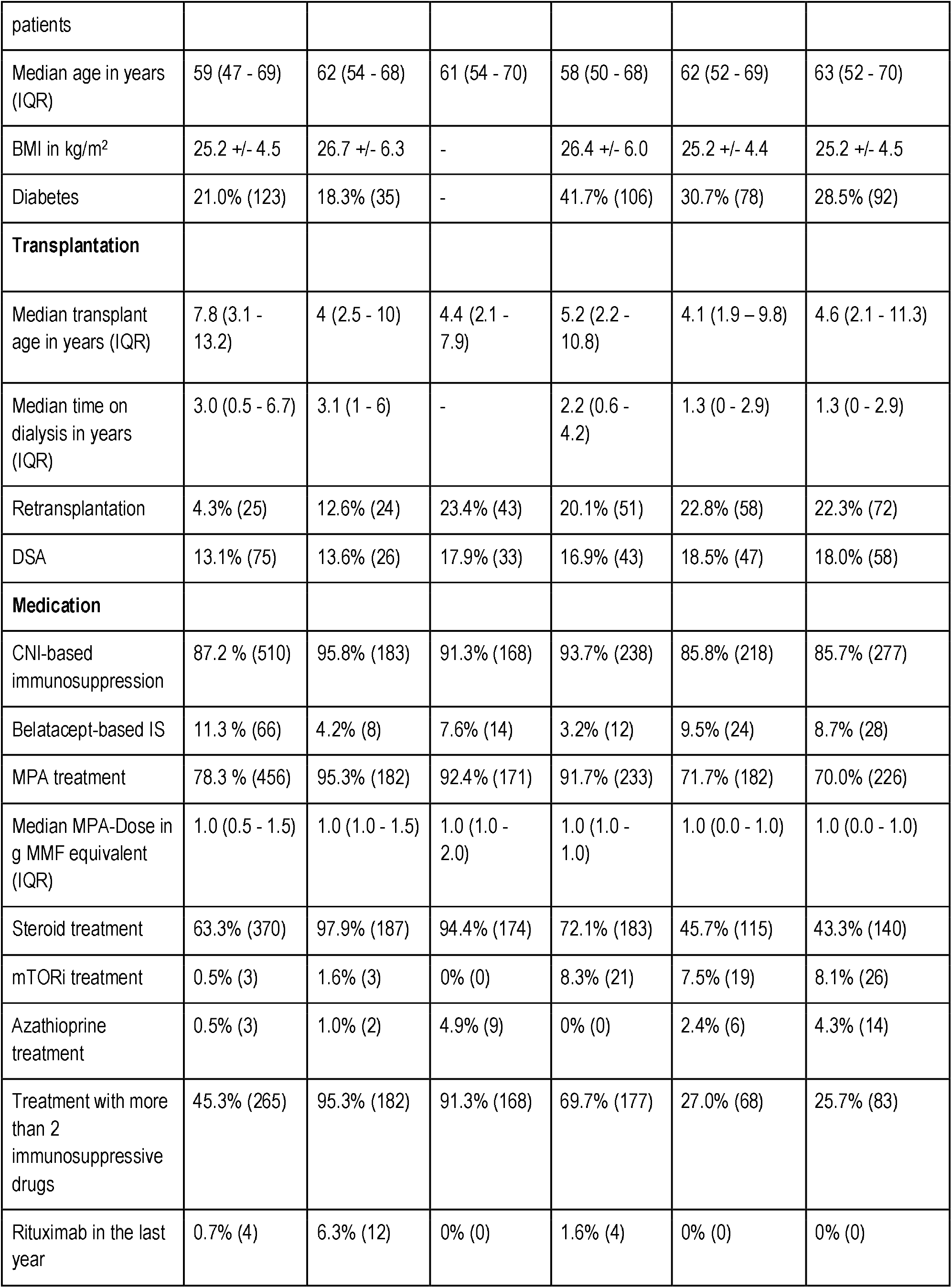
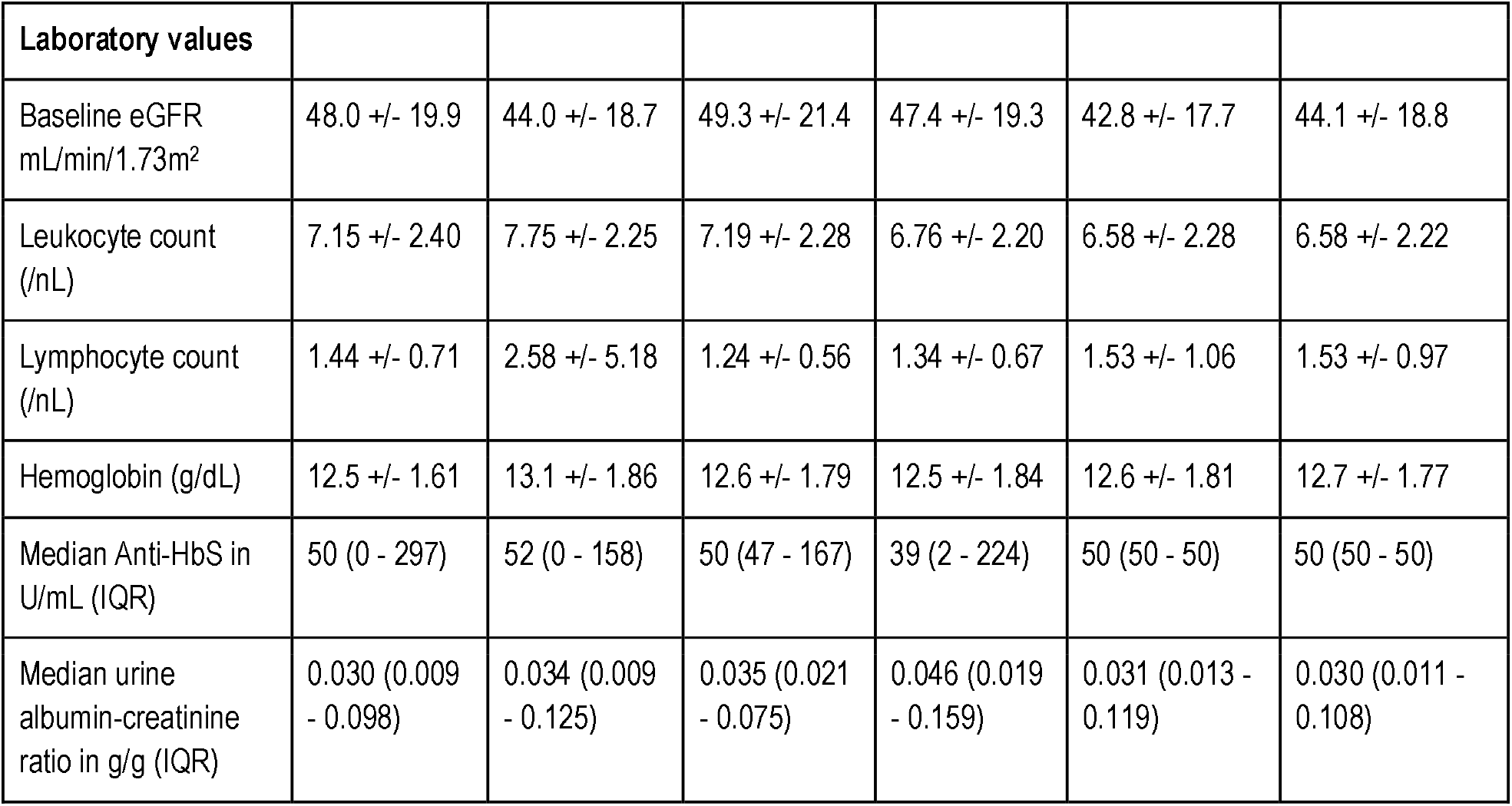
Baseline characteristics of the development and validation cohorts. All variables are reported as mean +/- standard deviation unless stated otherwise. IQR - interquartile range, mRNA – messenger ribonucleic acid, IgG – immunoglobulin G, BMI – body mass index, DSA – donor-specific anti human leukocyte antigen antibodies, CNI – calcineurin inhibitor, IS – immunosuppression, MPA – mycophenolic acid, MPA dose – mycophenolic acid dose, MMF – mycophenolic acid, mTORi – mammalian target of rapamycin inhibitor, eGFR – estimated glomerular filtration rate, anti-HBs – anti hepatitis B-surface-antigen immunoglobulin G antibodies.

### Development and internal validation

Using the development cohort, we evaluated five models during internal validation. To perform model validation within the development cohort, a resampling approach was used by assigning 585 vaccinations randomly 100 times into training and test sets of 410 and 175 each (70:30 split). Each time, hyperparameter tuning, if applicable, and model fitting was performed on the respective training set, and performance metrics were assessed on the respective test set.

First, as baseline, we fit a logistic regression model with all candidate variables using the R package *glm*.

Second, we fit 2 logistic regression models with least absolute shrinkage and selection operator (LASSO) regularization using the packages *caret* and *glmnet* in R. The LASSO hyperparameter λ, which adjusts the tradeoff between model fit and model sparsity, was optimized for each training cohort with respect to the mean squared error (MSE) using inner 5-fold cross-validation. We chose 2 different λ optimization criteria yielding 2 different models for each training cohort: (1) minimization of the MSE (termed LASSO-Min model), and (2) penalty maximization while keeping the MSE within one standard error of the minimum MSE (termed LASSO-1SE model).

Third, we fit a random forest regression model using the package *randomForest* in R. We optimized the hyperparameter mtry by evaluating 15 random parameter combinations during two repeated 5-fold cross-validations within the training set. The value of mtry yielding the highest accuracy during cross-validation was used to fit the random forest on the respective training data.

Fourth, we fit a gradient boosted regression trees (GBRT) model using the *gbm* package. We used a tune grid with 4*8*3*1 hyperparameter combinations (n.trees: 300, 500, 700, 900; interaction.depth: 2, 4, 6, 8, 10, 12, 14, 16; shrinkage: 0.001, 0.01, 0.1; n.minobsinnode: 10) to optimize hyperparameters during two repeated 5-fold cross-validations within the training set. The combination yielding the highest normalized discounted cumulative gain during cross-validation was used to fit the GBRT on the respective training data.

We calculated median performance during resampling for those five developed models. To evaluate the performance of the binary classification, we used Area Under the Curve of the Receiver Operator Characteristic (AUC-ROC), and confidence intervals (CI) in the resampling approach were determined from the empirical 2.5% and 97.5% quantiles of the performance on the 100 different test sets. Based on the threshold determined by the optimization criterion “closest.topleft” as provided in R package pROC (point with the least distance to [0,1] on the ROC-curve) during ROC-analysis, we calculated models’ sensitivity, specificity, accuracy, positive predictive value, and negative predictive value for each resampling step, again yielding median and empirical 95% CI.

### External validation and implementation

Due to their performance, we chose LASSO-Min and LASSO-1SE for estimation of model coefficients in the entire development cohort, which were then used for external validation. The relationship between the hyperparameter λ that controls model sparsity and the MSE during inner 5-fold cross-validation is shown in **Figure S6**. We assessed the thresholds for classification by determining the “closest.topleft” threshold on the entire development cohort, which were used for classification during external validation and are also provided in the online risk calculator after transforming into risk probability according to the formula:

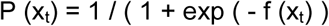

where *x*_*t*_ is the risk score threshold.

For external validation, we calculated the aforementioned performance metrics on each validation cohort separately. Furthermore, 95% CIs in the external validation cohorts were determined by performing 1000-fold ordinary nonparametric percentile bootstrap, as the empirical 2.5%, and 97.5% quantiles of AUC, sensitivity, specificity, accuracy, positive predictive value, and negative predictive value based on the thresholds determined within the development cohort.

To make the prediction model publicly available, we created an online tool implementing the models used for external validation, which can be assessed at https://www.tx-vaccine.com. Statistical analysis was performed using R studio v.1.2.5042 and R version 4.1.2 (2021-11-01). The underlying code is available on request from the corresponding author.

This article was prepared according to the transparent reporting of a multivariable prediction model for individual prognosis or diagnosis (TRIPOD) statement and we provided a checklist in the supplement.^18^

## Results

In total, 585 vaccinations (407 third vaccinations, and 178 fourth vaccinations) were used for development and internal validation, which is summarized together with outcome frequencies and reasons for exclusion in **Figure 1**.

Baseline characteristics of patients in the development and validation datasets including summary statistics of all variables are shown in **Table 3**.

### Model development and internal validation

Using the resampling approach outlined above, we fit five different models on each training set and evaluated their performance on the respective unseen test set during 100 resampling steps.

A logistic regression model employing all candidate variables served as a baseline. Using the two different λ optimization criteria outlined in “Methods”, the LASSO-Min and LASSO-1SE models were fitted. Additionally, two tree-based machine-learning approaches were studied -random forest (RF) and gradient boosted regression trees (GBRT).

LASSO logistic regression selected in the majority of resampling runs 23 and 11 out of 27 potential predictors to yield the LASSO-Min and LASSO-1SE models, respectively. The regression coefficients, their variances, and the selection frequency of the predictors are shown in **Figure S7 and S8**.

**Figure 2** compares AUC-ROC of the 5 models on the unseen test sets during 100 resampling steps, and **Table 4** summarizes median performance metrics as well as 95% confidence intervals determined from empirical 2.5% and 97.5% quantiles during internal validation. Thresholds for binary classification were determined on the respective test set during each resampling step by performing ROC-analysis.

**Figure 2.**
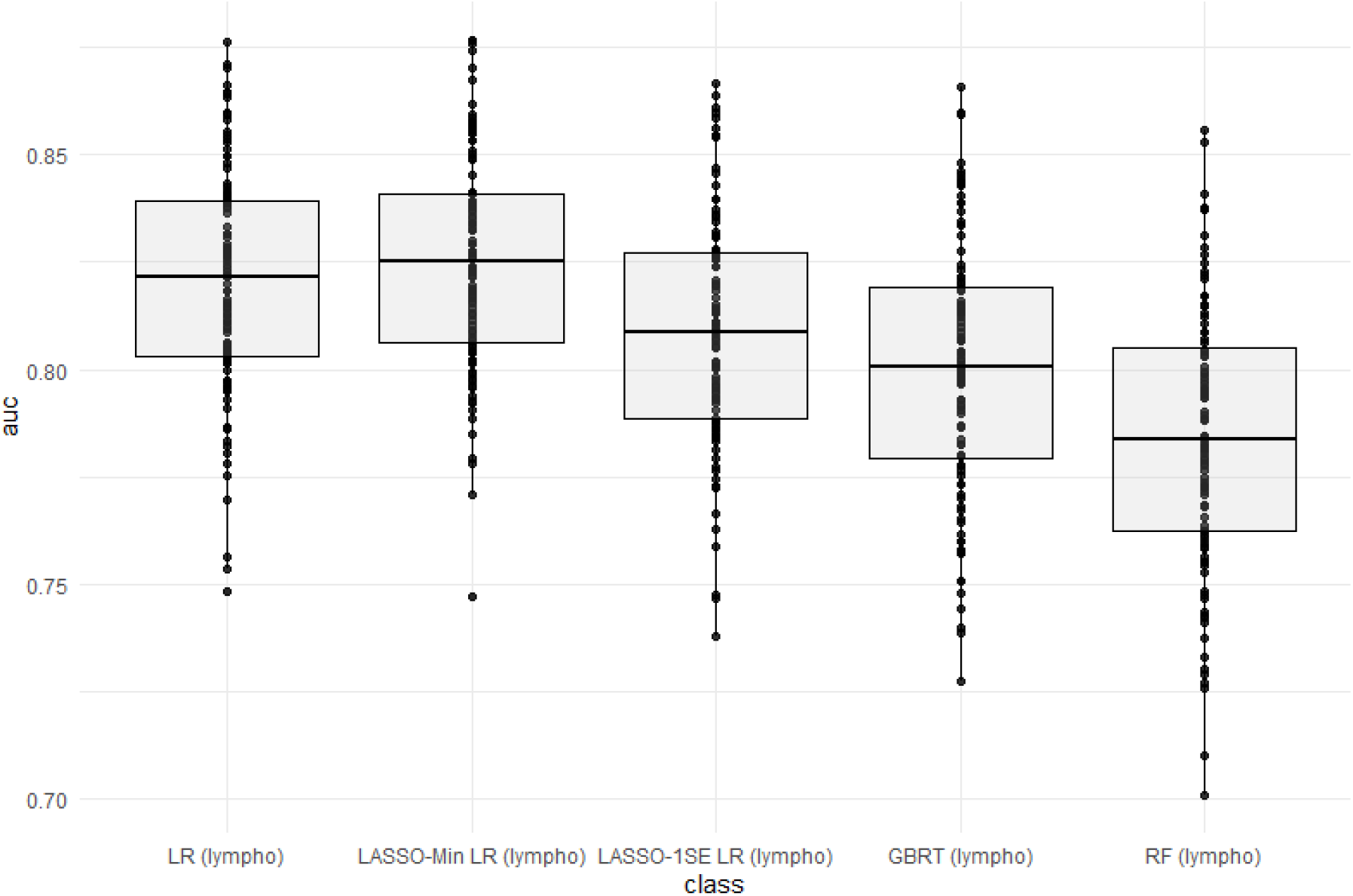
Predictive performance of the developed models (AUC) in internal validation. LR - logistic regression, LASSO-Min LR - least absolute shrinkage and selection operator regularized logistic regression with lambda hyperparameter optimized to yield minimum mean squared error within an inner 5-fold cross validation in the training set. LASSO-1SE - least absolute shrinkage and selection operator regularized logistic regression with lambda hyperparameter increased from lambda-min, so that mean squared error stays within one standard error within an inner 5-fold cross validation in the training set. GBRT - gradient boosted regression trees. RF - random forest. lympho – including lymphocyte count as predictor variable.

**Table 4.** Performance of five different models during internal validation. AUC-ROC, as well as sensitivity (Sens), specificity (Spec), accuracy (Acc.), positive predictive value (PPV), negative predictive value (NPV) in the test set based on the best threshold during ROC-analysis. Median and empirical 95% CI are derived from 100 resampling steps for each metric.

| Model Type | AUC | Sens | Spec | Acc | PPV | NPV |
| --- | --- | --- | --- | --- | --- | --- |
| Logistic Regression | 0.822<br>(0.763 - 0.868) | 0.763<br>(0.690 - 0.838) | 0.784<br>(0.679 - 0.861) | 0.778<br>(0.707 - 0.827) | 0.671<br>(0.563 - 0.764) | 0.851<br>(0.795 - 0.898) |
| LASSO-Min | 0.825<br>(0.779 - 0.875) | 0.767<br>(0.679 - 0.863) | 0.773<br>(0.685 - 0.859) | 0.776<br>(0.712 - 0.821) | 0.667<br>(0.546 - 0.767) | 0.854<br>(0.797 - 0.907) |
| LASSO-1SE | 0.809<br>(0.753 - 0.860) | 0.746<br>(0.654 - 0.835) | 0.761<br>(0.681 - 0.844) | 0.756<br>(0.696 - 0.813) | 0.649<br>(0.533 - 0.743) | 0.833<br>(0.778 - 0.896) |
| Random Forest | 0.784<br>(0.726 - 0.839) | 0.724<br>(0.620 - 0.817) | 0.762<br>(0.646 - 0.848) | 0.750<br>(0.673 - 0.795) | 0.634<br>(0.503 - 0.739) | 0.825<br>(0.763 - 0.894) |
| GBM | 0.800<br>(0.742 -<br>0.854) | 0.741<br>(0.626 -<br>0.842) | 0.757<br>(0.652 -<br>0.862) | 0.750<br>(0.688 -<br>0.804) | 0.642<br>(0.519 -<br>0.763) | 0.831<br>(0.775 -<br>0.898) |

With respect to AUC-ROC, the LASSO-Min model - 0.825 (0.779 - 0.875) and the baseline logistic regression model - 0.822 (0.763 - 0.868) showed best performance during internal validation. Since the sparser LASSO-1SE model showed comparable predictive performance of 0.809 (0.753 - 0.860) with fewer variables, we chose to use both, LASSO-Min and LASSO-1SE regularized logistic regression models for external validation. Due to higher model complexity with comparably worse predictive performance, we did not follow RF and GBRT for external validation.

### Model specification

Final risk equations were obtained by fitting LASSO-Min and LASSO-1SE models on the complete development dataset, yielding a 23-variable and 11-variable risk equation, respectively. The intercept and regression coefficients of the final models are shown in **Table 5**. Risk equations are provided in **Items S5 and S6**, and are implemented as an online tool available at https://www.tx-vaccine.com.

**Table 5.**
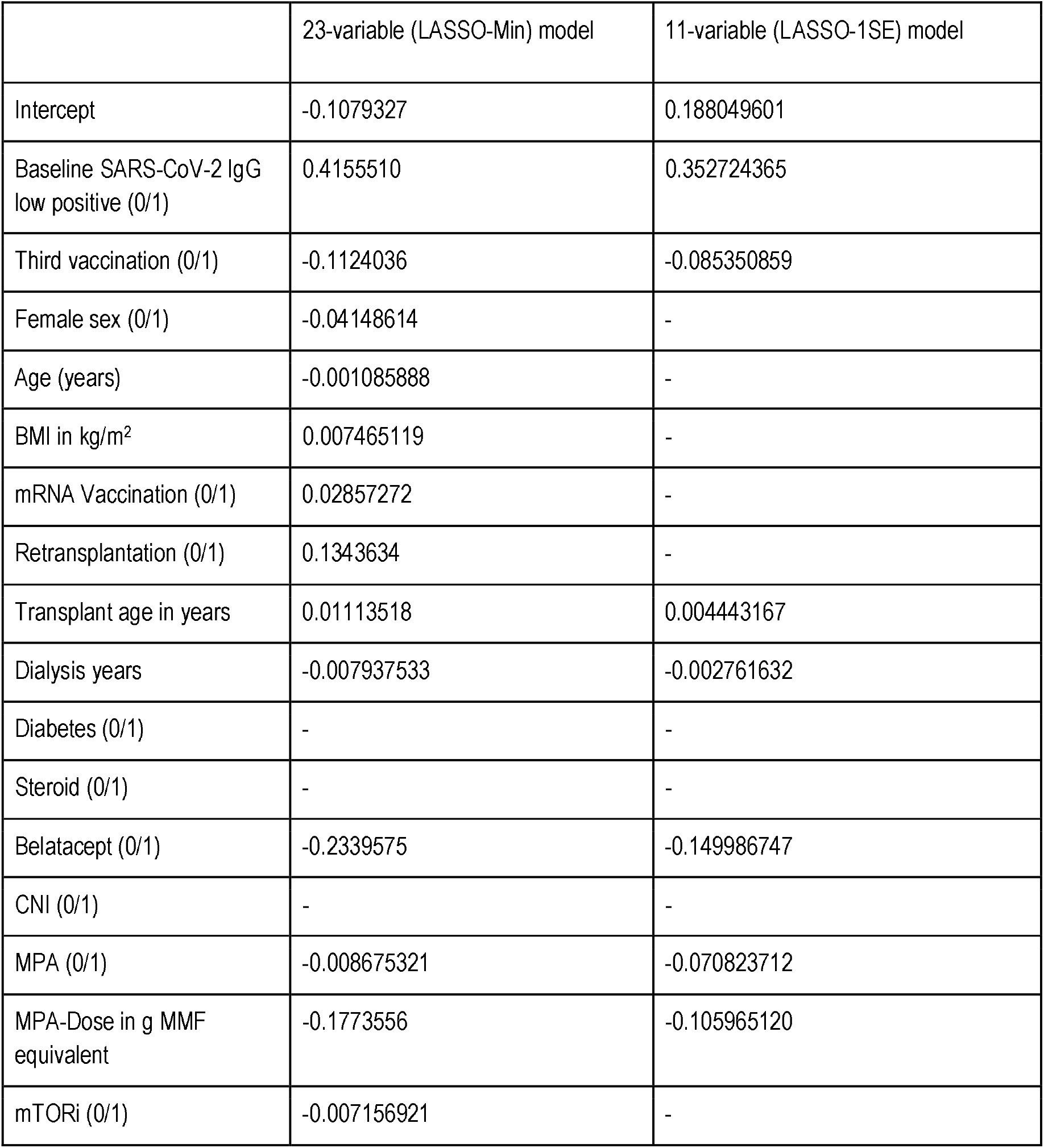
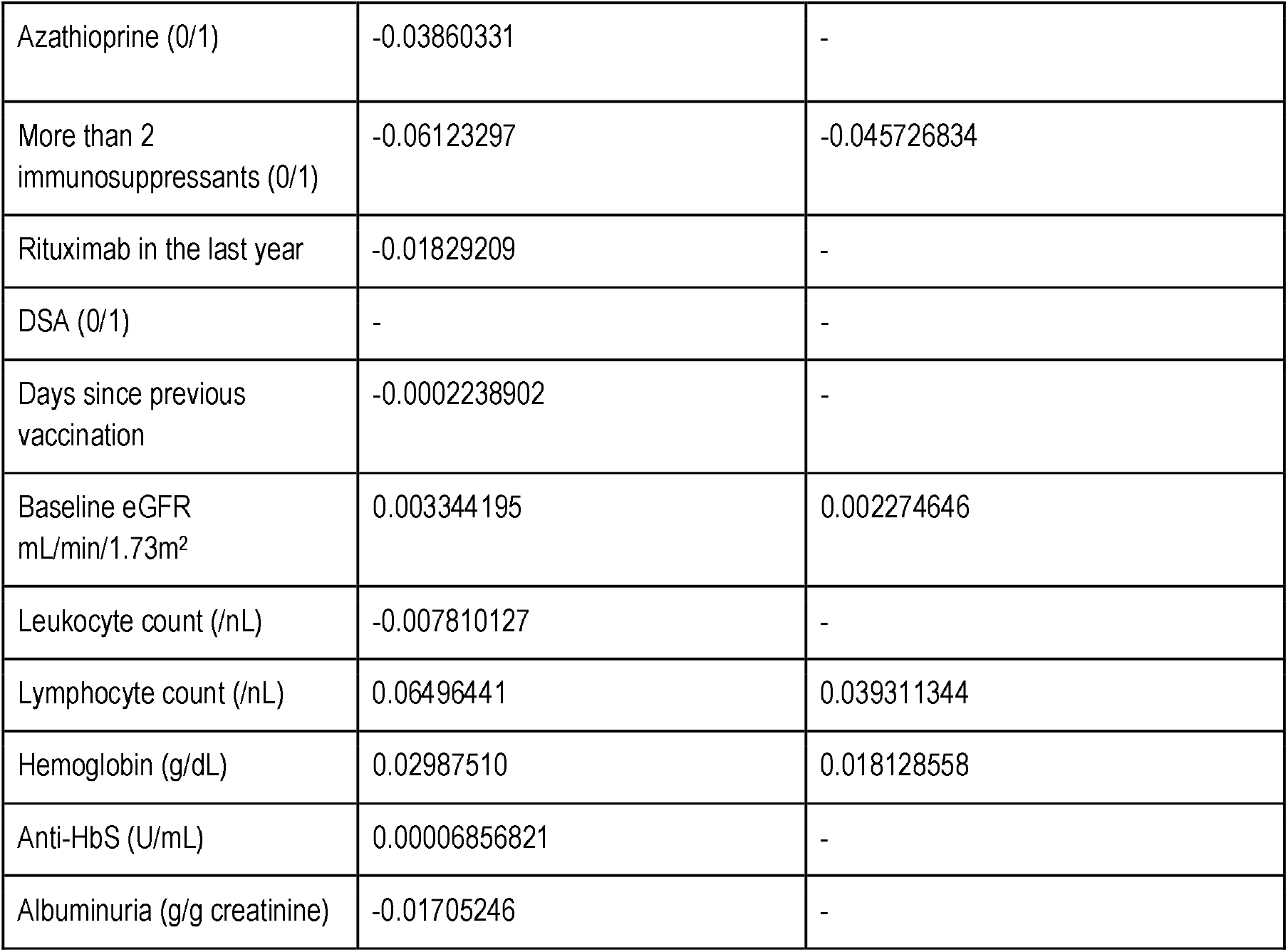
Final intercept and coefficients of the 23-variable (LASSO-Min), and 11-variable (LASSO-1SE) logistic regression model fitted on the entire development dataset, both of which are used for external validation.

|  | 23-variable (LASSO-Min) model | 11-variable (LASSO-1SE) model |
| --- | --- | --- |
| Intercept | -0.1079327 | 0.188049601 |
| Baseline SARS-CoV-2 IgG low positive (0/1) | 0.4155510 | 0.352724365 |
| Third vaccination (0/1) | -0.1124036 | -0.085350859 |
| Female sex (0/1) | -0.04148614 | - |
| Age (years) | -0.001085888 | - |
| BMI in kg/m<sup>2</sup> | 0.007465119 | - |
| mRNA Vaccination (0/1) | 0.02857272 | - |
| Retransplantation (0/1) | 0.1343634 | - |
| Transplant age in years | 0.01113518 | 0.004443167 |
| Dialysis years | -0.007937533 | -0.002761632 |
| Diabetes (0/1) | - | - |
| Steroid (0/1) | - | - |
| Belatacept (0/1) | -0.2339575 | -0.149986747 |
| CNI (0/1) | - | - |
| MPA (0/1) | -0.008675321 | -0.070823712 |
| MPA-Dose in g MMF equivalent | -0.1773556 | -0.105965120 |
| mTORi (0/1) | -0.007156921 | - |
| Azathioprine (0/1) | -0.03860331 | - |
| More than 2 immunosuppressants (0/1) | -0.06123297 | -0.045726834 |
| Rituximab in the last year | -0.01829209 | - |
| DSA (0/1) | - | - |
| Days since previous vaccination | -0.0002238902 | - |
| Baseline eGFR mL/min/1.73m<sup>2</sup> | 0.003344195 | 0.002274646 |
| Leukocyte count (/nL) | -0.007810127 | - |
| Lymphocyte count (/nL) | 0.06496441 | 0.039311344 |
| Hemoglobin (g/dL) | 0.02987510 | 0.018128558 |
| Anti-HbS (U/mL) | 0.00006856821 | - |
| Albuminuria (g/g creatinine) | -0.01705246 | - |

### External validation

After applying all exclusion criteria and performing imputation of missing variables, we evaluated both risk equations in the four independent validation datasets. Since predictive performance during external validation was comparable for both models, in the following we report on the sparser 11-variable model. Results of external validation of the 23-variable model are summarized in **Table S3**, and **Figure S9**.

AUC-ROC of the sparser 11-variable model during external validation was 0.855 (0.799 - 0.911) for validation set 1, 0.722 (0.647 - 0.786) for validation set 2, 0.828 (0.772 - 0.877) for validation set 3, and 0.708 (0.643 - 0.773) for validation set 4, yielding an AUC-ROC of 0.764 (0.732 - 0.795) when merging all validation sets. Sensitivity, specificity, accuracy, positive predictive value, and negative predictive value using the thresholds determined during ROC-analysis in the development dataset are summarized in **Table 6**. The decision thresholds used for external validation are also provided in the online risk calculator to guide physicians’ decision.

**Table 6.** Performance of the 11-variable model during external validation. AUC-ROC, as well as sensitivity (Sens), specificity (Spec), accuracy (Acc.), positive predictive value (PPV), negative predictive value assessed on each validation set. The threshold was derived during ROC-analysis on the development dataset. To provide 95% CI, empirical 2.5% and 97.5% quantiles of the respective metric are provided after performing a 1000-fold nonparametric ordinary bootstrapping with each validation set.

| Model Type | AUC point estimate (95% CI) | Sens point estimate (95% CI) | Spec point estimate (95% CI) | Acc point estimate (95% CI) | PPV point estimate (95% CI) | NPV point estimate (95% CI) |
| --- | --- | --- | --- | --- | --- | --- |
| Validation 1 11-variable | 0.855 (0.799 - 0.911) | 0.800 (0.700 - 0.891) | 0.674 (0.586 - 0.756) | 0.717 (0.649 - 0.780) | 0.559 (0.460 - 0.660) | 0.867 (0.796 - 0.927) |
| Validation 2 11-variable (cutoff 0.8/mL) | 0.722 (0.647 - 0.786) | 0.174 (0.088 - 0.278) | 0.922 (0.867 - 0.964) | 0.641 (0.565 - 0.707) | 0.571 (0.350 - 0.769) | 0.650 (0.570 - 0.719) |
| Validation 2<br>11-variable<br>(cutoff 15 U/mL) | 0.749<br>(0.672 -<br>0.819) | 0.205<br>(0.077 -<br>0.344) | 0.910<br>(0.863 -<br>0.952) | 0.761<br>(0.696 -<br>0.821) | 0.381<br>(0.167 -<br>0.611) | 0.810<br>(0.744 -<br>0.868) |
| Validation 3<br>11-variable | 0.828<br>(0.772 -<br>0.877) | 0.718<br>(0.637 -<br>0.790) | 0.738<br>(0.669 -<br>0.809) | 0.728<br>(0.669 -<br>0.800) | 0.724<br>(0.646 -<br>0.803) | 0.733<br>(0.656 -<br>0.801) |
| Validation 4<br>11-variable<br>(cutoff 0.8/mL) | 0.708<br>(0.643 -<br>0.773) | 0.667<br>(0.588 -<br>0.745) | 0.615<br>(0.522 -<br>0.706) | 0.646<br>(0.587 -<br>0.705) | 0.714<br>(0.640 -<br>0.784) | 0.561<br>(0.475 -<br>0.655) |
| Validation 4<br>11-variable<br>(cutoff 15 U/mL) | 0.787<br>(0.737 -<br>0.850) | 0.795<br>(0.735 -<br>0.850) | 0.585<br>(0.496 -<br>0.670) | 0.715<br>(0.666 -<br>0.762) | 0.757<br>(0.696 -<br>0.814) | 0.637<br>(0.542 -<br>0.725) |
| **Overall**<br>**performance**<br>**11-variable** | **0.764**<br>**(0.732 -**<br>**0.795)** | **0.620**<br>**(0.571 -**<br>**0.673)** | **0.739**<br>**(0.700 -**<br>**0.774)** | **0.684**<br>**(0.653 -**<br>**0.715)** | **0.671**<br>**(0.621 -**<br>**0.716)** | **0.694**<br>**(0.651 -**<br>**0.733)** |
| **Overall**<br>**performance**<br>**11-variable**<br>**(cutoff 15 U/mL)** | **0.819**<br>**(0.793 -**<br>**0.844)** | **0.720**<br>**(0.676 -**<br>**0.762)** | **0.735**<br>**(0.698 -**<br>**0.775)** | **0.728**<br>**(0.702 -**<br>**0.756)** | **0.689**<br>**(0.647 -**<br>**0.731)** | **0.762**<br>**(0.728 -**<br>**0.800)** |

To estimate confidence intervals of predictive performance metrics, we performed 1000 nonparametric ordinary bootstrapping for each validation set and assessed the same metrics as above. Median, as well as 95% confidence intervals derived from the empirical 2.5% and 97.5% quantiles are provided in **Figure 3** and **Table 6**.

**Figure 3.**
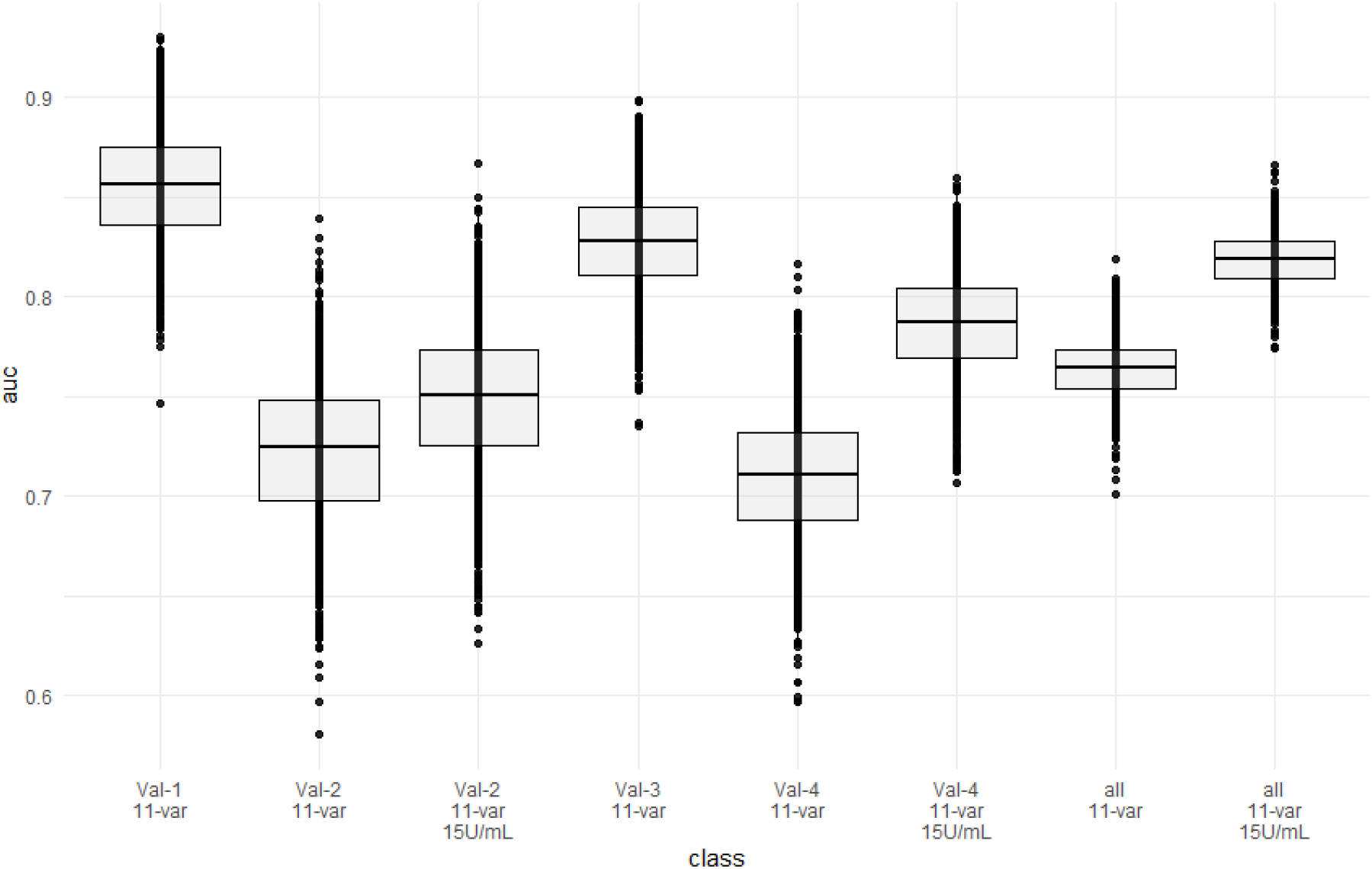
Predictive performance (AUC-ROC) of the 11-variable model in external validation.

### Implementation and cutoff definition

Performance in the validations sets 2 and 4 was poorer than in the development as well as in the other two validation sets. We suspected the positivity cutoff of 0.8 U/mL provided by the manufacturer for the ECLIA Elecsys assay as one main reason. Since it is close to the limit of detection (0.4 U/mL), no or small fraction of “low positive” antibody levels (values above the limit of detection and below positivity cutoff) before vaccination are present in both validations sets (**Table 3**), which is different to both other validation sets and the development dataset. Since a low positive antibody level before vaccination is an important predictor of serological response (**Table 5**), we chose to adjust the positivity cutoff to 15 U/mL arbitrarily for two reasons. First, to test the hypothesis that cutoff definition is a reason for lower performance. Second, to provide data that an implementation of this model is feasible independent of the assay used. Our proposed implementation strategy for the prediction model is to identify patients, who will not respond to an additional vaccine dose despite changes in immunosuppression, and to offer those patients passive immunization (**Figure 4**). Hence, instead of using the positivity cutoff from the manufacturer, using any other cutoff below which no neutralization against omicron occurs, is compatible with this strategy under the circumstances of omicron-dominance. As described in “Methods”, we arbitrarily use an alternative positivity cutoff of 15 U/mL for this respective assay, since it has already been proposed by the manufacturer before.

**Figure 4.**
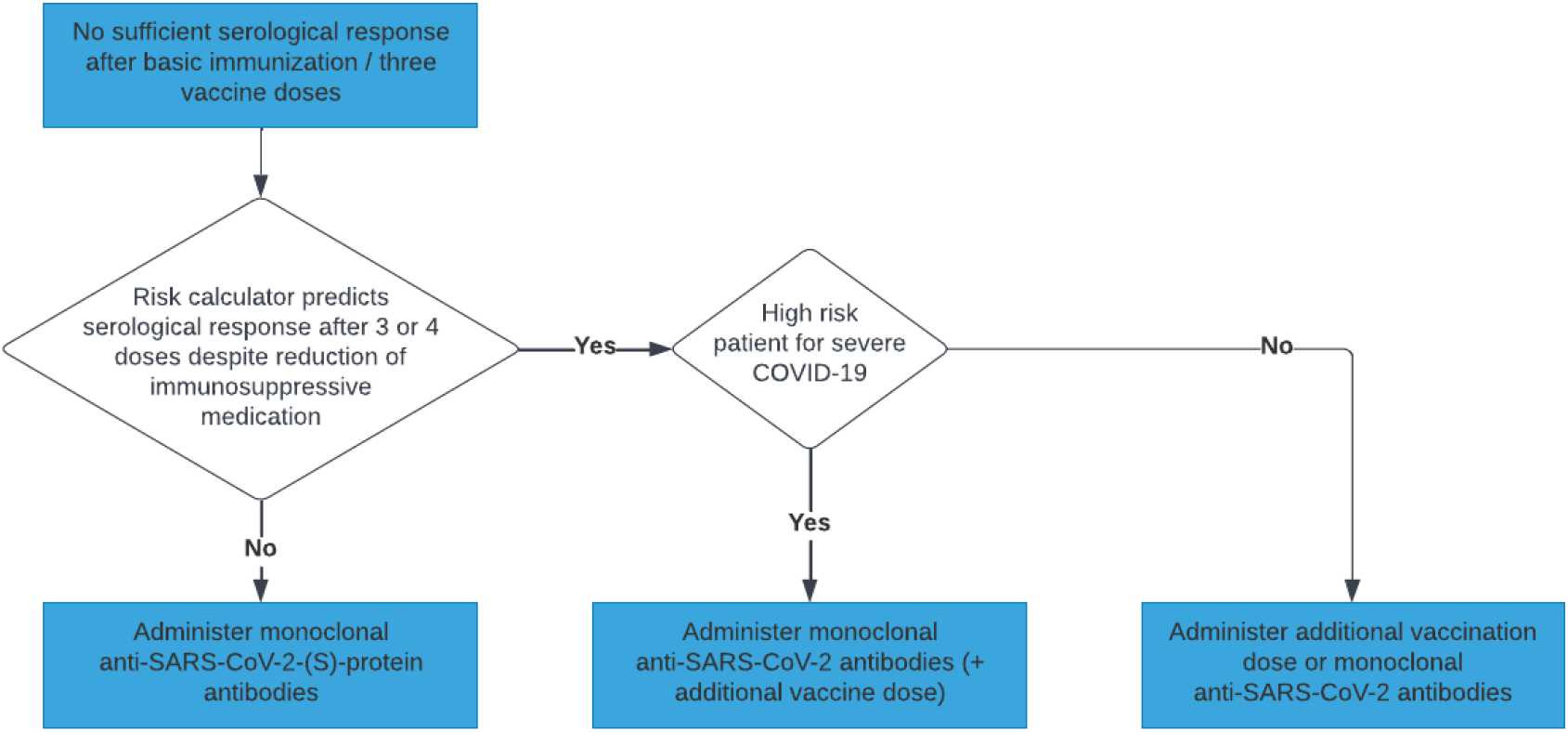
Possible implementation strategy of the described prediction model into clinical decision making.

When adjusting the assay cutoff to 15 U/mL for the ECLIA Elecsys assay, AUC-ROC increased to 0.749 (0.672 - 0.819) for validation set 2, and to 0.787 (0.737 - 0.850) for validation set 4, yielding an overall AUC-ROC of 0.819 (0.793 - 0.844) after merging all validation sets. With the thresholds assessed in the development dataset, the model achieves a negative predictive value of 0.76 (0.728 - 0.800).

## Discussion

In this article, we present the development, as well as internal and external validation of an 11-variable LASSO regularized logistic regression model for prediction of serological response to the third and fourth dose of SARS-CoV-2 vaccine in seronegative kidney transplant recipients.

It shows good discrimination of KTR exhibiting serological response both in a rigorous resampling approach in the development cohort and in four independent validation cohorts with positive and negative predictive values of 0.76 based on a threshold established within the development dataset. Available online as a risk calculator at https://www.tx-vaccine.com and embedded into the proposed implementation strategy, it can assist physicians in choosing between different immunization strategies, namely, additional vaccination with or without adaption of immunosuppressive therapy, or pre-exposure prophylaxis with monoclonal anti-SARS-CoV-2-(S) antibodies.

While serological response is only one part of the immune response to vaccination, neutralizing anti-SARS-CoV-2-(S) antibodies are pathophysiologically and epidemiologically established to offer protection from severe disease ^9,19^, which is also supported by the protection offered by monoclonal antibodies against SARS-CoV-2 applied for prophylaxis and treatment.^20,21^

Yet, after the emergence of the omicron variants, neutralization antibody levels against omicron variant show 25.7-fold to 58.1-fold reduction in sera of healthy vaccinated subjects in comparison to wild-type.^22^ Consequently, antibody levels that ensure neutralization, increased from >264 U/mL for alpha variant ^8,9^ to >2000 U/mL for omicron ^16^, making the interpretation of antibody levels more difficult than before.

Despite these uncertainties, it seems impossible to leave patients without any humoral protection whatsoever. Hence, in patients without serological response to basic immunization, physicians and patients need to decide between additional active vaccination with or without adapting immunosuppressive medication, and pre-exposure prophylaxis with monoclonal antibodies.^23^

Since negative predictive value was above 0.75, when merging all validation sets, and above 0.8 in the development dataset and in two out of four validation datasets, when using the higher positivity cutoff of 15 U/mL for the ECLIA Elecsys assay, we suggest the following implementation strategy summarized in **Figure 4**. For patients, who are likely not to respond to additional SARS-CoV-2 vaccination according to the prediction model, pre-exposure prophylaxis with monoclonal antibodies exhibiting neutralizing capacity against omicron BA.2 should be administered to ensure timely protection.^24-27^ In patients, who are likely to respond according to the prediction model, there is still a chance that these patients will not reach antibody levels, which ensure neutralizing capacity against omicron variants. For these patients, both, repeated vaccination and monoclonal antibody prophylaxis are feasible and should in our view be chosen depending on the risk for severe disease course. When evidence about antibody levels that ensure sufficient neutralization against omicron increases, an updated model for the prediction of high-responders that do not warrant monoclonal antibody therapy can be made available.

## Strengths and Limitations

We provide a rigorously developed and validated prediction model, which is provided as an online risk calculator to support kidney transplant physicians in their daily practice when deciding upon the immunization strategy for their patients. Especially, the estimated effects of adaptions in immunosuppressive medication can be evaluated, e.g. reducing or pausing MPA dose, or switching from belatacept to calcineurin inhibitor. Regarding the general sparsity of data on vaccine response to third and fourth dose in KTR, we analyze extensive datasets for development and validation, and hereby provide representative evaluation of real-life model performance. While performance of the 23-variable model was slightly better, it is also more impractical due to the high number of variables, which is why we chose to report mainly on the sparser 11-variable model.

In general, our model detects patients who are likely to respond or not respond at all to an additional vaccine dose, and is able to factor in changes in immunosuppressive medication, when making individual predictions. Since evidence of antibody level cutoffs that ensure neutralization of or protection from omicron is sparse, we chose not to make any predictions for this endpoint. Instead, we provide an implementation strategy that makes best use of the model’s prediction without making far-reaching assumptions about protective antibody levels against omicron.

Still, one major limitation becomes evident for validation sets 2 and 4, where predictive performance was only moderate when using the positivity cutoff of 0.8 U/mL for the ECLIA Elecsys assay. As outlined above, increasing the positivity cutoff to 15 U/mL is compatible with the proposed implementation, and leads to improved performance for two reasons.

First, since the cutoffs in the development dataset were based on protective levels against alpha variant for the ECLIA Elecsys assay (264 U/mL), predictive performance is expectably poorer when predicting positivity with a 0.8 U/mL cutoff. Second, with a cutoff of 0.8 U/mL for the ECLIA Elecsys assay, only very few patients with low positive antibody levels pre vaccination exist, with low positive being every value above zero or limit of detection (LoD), but below the positivity cutoff. Since this is an important predictor in both models and provides important information about the actual immunological status, loss of performance can be expected when this information is missing. This is further supported by the fact that after adapting the cutoff from 0.8 U/mL to 15 U/mL, in validation set 2, where all pre-vaccination antibody levels were below 0.4 U/mL, the performance only increased slightly (AUC-ROC 0.722 to 0.749), but in validation set 4, where the percentage of low positive patients increased from 14.6% (most of which were due to the other assays used in this dataset) to 33.1%, the performance increased markedly (AUC-ROC 0.708 to 0.787).

Other possible reasons for different performance are the study design of validation set 2, which was a randomized clinical trial, with outcome assessment between days 29 and 42, whereas in the development and other validation sets, the maximum antibody level after the respective vaccination was chosen, independent of the time passed. Additionally, validation set 2 was the one with the highest proportion of adenoviral vaccine, which however, did not show any difference in serological response in the respective trial.^12^ The remaining 3 validation sets, were mostly assessing serological response to guide further vaccine doses, similarly to the development cohort.

Worth noticing is the imputation method used, which ensures that performance assessed during external validation is comparable to real-life performance of the risk calculator provided.

Other limitations arise from the different immunization strategies used, which lead to different seroconversion rates and have influence on model performance as well.

Last, some candidate variables have low frequency below 1% in the development dataset (such as rituximab in the last year, mTOR inhibitor treatment, azathioprine treatment), which limits generalizability for these patients.

## Conclusion

We provide the first, online available calculator to predict vaccine response to third or fourth vaccination in kidney transplant recipients together with a clear implementation strategy that enables physicians to substantiate their decision making with respect to the optimal immunization strategy.

## Data Availability

All data produced in the present study are available upon reasonable request to the authors.

## Acknowledgements

Eva Schrezenmeier is a participant in the BIH-Charité Clinician Scientist Program funded by the Charité–Universitätsmedizin Berlin and the Berlin Institute of Health.

We thank Jürgen Dönitz, Johannes Raffler, Ulla Schultheiss, and Helena U. Zacharias on behalf of the CKDapp team for providing their template for an online risk calculator.

## Supplemental Table of Contents

**Table S1**. Variable definition for descriptive statistics and multivariable analysis.

**Item S1**. Detailed information about validation cohort 1

**Figure S1**. Patient flow diagram of the validation cohort 1.

**Item S2**. Detailed information about validation cohort 2

**Figure S2**. Patient flow diagram of the validation cohort 2.

**Item S3**. Detailed information about validation cohort 3.

**Figure S3**. Patient flow diagram of the validation cohort 3.

**Item S4**. Detailed information about validation cohort 4

**Figure S4**. Patient flow diagram of the validation cohort 4.

**Figure S5**. Imputation or including patients without lymphocyte count did not achieve better performance during internal validation than complete case analysis.

**Table S2**. Preliminary analysis did not show improvement of predictive accuracy of logistic regression models when excluding lymphocyte count as a predictor variable or by performing multiple imputation for missing laboratory values.

**Figure S6**. Relationship between model sparsity and predictive performance for the LASSO logistic regression on the development dataset.

**Figure S7**. Estimated LASSO coefficients of the LASSO-min models summarized across 100 subsampling runs for unstandardized variables.

**Figure S8**. Estimated LASSO coefficients of the LASSO-1SE risk scores summarized across 100 subsampling runs for unstandardized variables.

**Table S3**. Performance of the 23-variable model during external validation.

**Figure S9**. Predictive performance of both the 23-variable and 11-variable models in four independent external validation cohorts employing two different cutoff definitions for ECLIA Elecsys antibody assay of 0.8 U/mL and 15 U/mL, respectively.

