## Supplementary Data for "Development and Validation of Multivariable Prediction Models of Serological Response to SARS-CoV-2 Vaccination in Kidney Transplant Recipients"

**Table S1.** Variable definition for descriptive statistics and multivariable analysis.

| **Candidate Variable** | **Data Type** | **Definition** |
| --- | --- | --- |
| Third vaccination | binary (1/0) | Third (not fourth) SARS-CoV-2 vaccine dose in the respective patients |
| mRNA vaccine | binary (1/0) | respective vaccination performed with an mRNA-based SARS-CoV-2 vaccine |
| Time since last vaccination | integer | time passed since the last SARS-CoV-2 vaccination in days |
| Baseline SARS-CoV-2 IgG low positive | binary (1/0) | SARS-CoV-2 antibody levels above 0 or the limit of detection, but below the positivity cutoff before the respective vaccination |
| Female sex | binary (1/0) | biological sex (female=1 or male=0) |
| Age | float | age at the time of vaccination in years |
| BMI | float | body mass index (body mass divided by square of the body height) |
| Diabetes | binary (1/0) | a diagnosis of diabetes mellitus in the patient's history or current use of antidiabetic medication |
| Transplant age | float | time since the patient's last kidney transplantation at the time of vaccination in years |
| Retransplantation | binary (1/0) | Current kidney transplant is not the first kidney transplant for the respective patient |
| Dialysis time | float | Cumulative time on dialysis in years |
| DSA | binary (1/0) | Presence of donor-specific anti-human leukocyte antigen antibodies |
| CNI | binary (1/0) | The use of systemic tacrolimus or cyclosporine as an immunosuppressive medication at the time of vaccination |
| Belatacept | binary (1/0) | The use of belatacept as an immunosuppressive medication at the time of vaccination |
| MPA | binary (1/0) | The use of mycophenolic acid (as mycophenolate sodium or mycophenolate mofetil (MMF)) as an immunosuppressive medication at the time of vaccination |
| MPA dose | float | mycophenolic acid dose in MMF equivalent in g/day at the time of vaccination |
| Steroid | binary (1/0) | The use of systemic steroids as an immunosuppressive medication at the time of vaccination |
| mTORi | binary (1/0) | The use of systemic sirolimus or everolimus as an immunosuppressive medication at the time of vaccination |
| Azathioprine | binary (1/0) | The use of systemic azathioprine as an immunosuppressive medication at the time of vaccination |
| More than 2 immunosuppressive drugs | binary (1/0) | The use of more than two types of immunosuppressive medication at the time of vaccination |
| RTX in the last year | binary (1/0) | administration of rituximab in the last 365 days before the respective vaccination |
| eGFR | float | most recent estimated glomerular filtration rate according to the Chronic Kidney Disease Epidemiology Collaboration (CKD-EPI) equation in ml/min/1.73m² at the time of vaccination (if unavailable, first measurement after vaccination) |
| White blood cell count | float | most recent white blood cell count in /nL at the time of vaccination (if unavailable, first measurement after vaccination) |
| Lymphocyte | float | most recent lymphocyte count in /nL at the time of vaccination (if unavailable, first measurement after vaccination) |
| Hemoglobin | float | most recent hemoglobin level in mg/dL at the time of vaccination (if unavailable, first measurement after vaccination) |
| anti-HBs | float | most recent presence/level of Hepatitis B surface antibodies at the time of vaccination (if unavailable, first measurement after vaccination) |
| Urine albumin creatinine ratio | float | most recent urinary albumin-to-creatinine ratio (ACR) in either spot or  24 hour collection urine in g/g at the time of vaccination (if unavailable, first measurement after vaccination) |

**Item S1.** Detailed information about validation cohort 1

The first validation dataset comprised of 239 vaccinations from the kidney transplant center at the University Hospital Düsseldorf, which were derived from a prospective observational trial. All patients provided written informed consent and institutional review board approval was given (ID 2020-1237) for the respective study. After applying all exclusion criteria, 191 vaccinations were included into the final analysis.

**Outcome and Predictors**

The SARS-CoV-2 IgG assay used was EUROIMMUN Anti-SARS-CoV-2 QuantiVac ELISA IgG Assay with BAU/mL as a readout. According to the manufacturer, a value ≥ 35.2 BAU/mL was considered positive. Non-zero values below the cutoff were considered low positive.

Remaining candidate variables were defined similarly to the development dataset.

**Handling of missing values**

When performing complete case analysis, only 127 vaccinations remained, which had no missing data for all 27 candidate predictors. In case of missing BMI, but available weight data, we imputed height as follows: 1.75 m for male patients, and 1.65 m for female patients.

To assess the practical utility of the risk calculator, we chose the imputation method outlined in methods. Namely, we imputed the median value of the development dataset, since this is the way the online risk calculator handles missing values. Reasons for exclusion and outcome frequencies are summarized in **Figure S1**.

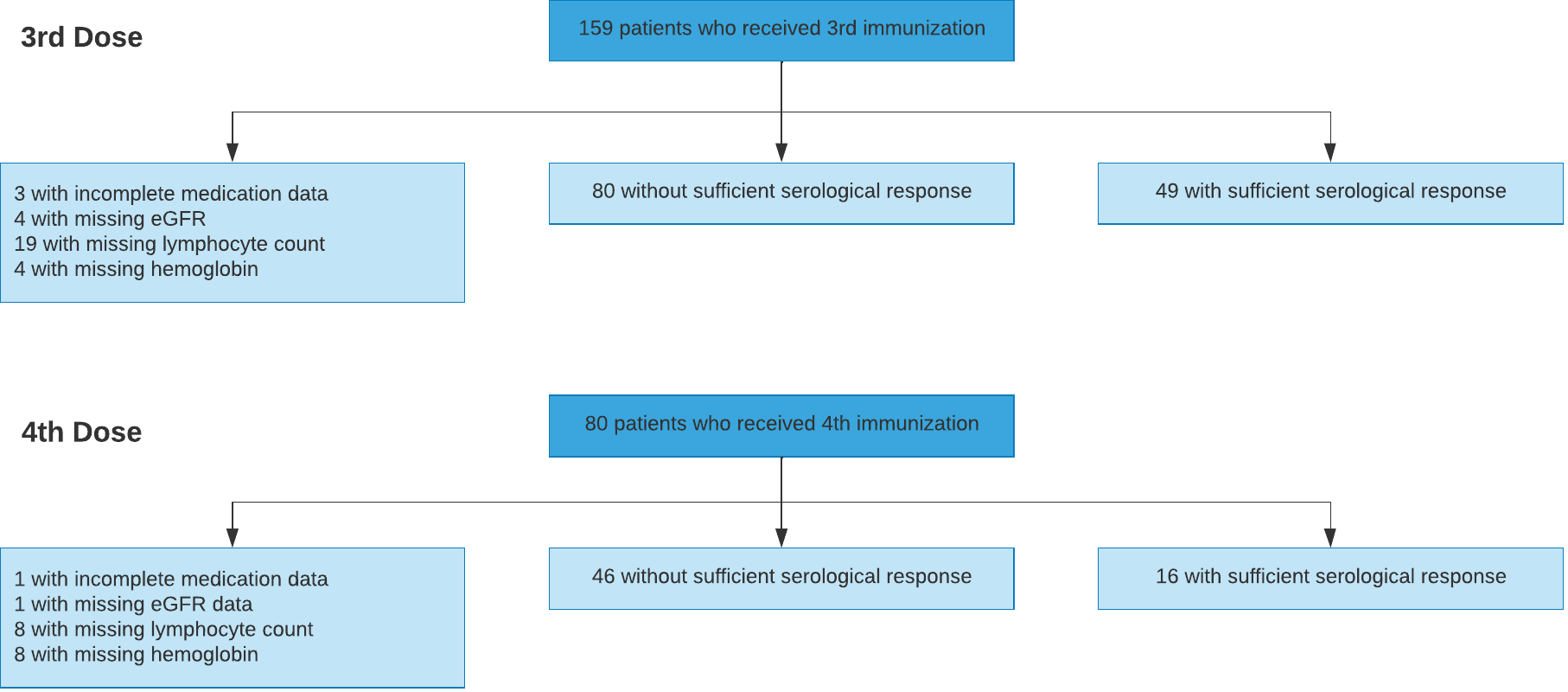

**Figure S1**. Patient flow diagram of the validation cohort 1.

**Item S2.** Detailed information about validation cohort 2

The second validation dataset consisted of 197 vaccinations from the kidney transplant center at the Medical University of Vienna (Austria), which were derived from a single center, single-blinded, 1:1 randomized clinical trial. Briefly, patients who had not developed SARS-CoV-2 spike protein antibodies after 2 doses of an mRNA vaccine, received either a third dose of the previously administered mRNA vaccine (mRNA-1273 or BNT162b2) or a single dose of the vector vaccine (Ad26COVS1). The trial was registered with the European Union Clinical Trial Register and was approved by the ethics committee of the Medical University of Vienna (No. 1612/2021) as well as by the Austrian Agency for Health and Food Safety.

**Outcome and Predictors**

The SARS-CoV-2 assay used was the electrochemiluminescence immunoassay (ECLIA) (Elecsys, Anti-SARS-CoV-2, Roche Diagnostics GmbH, Mannheim, Germany) with BAU/mL as a readout. According to the manufacturer, a value above 0.8 U/mL was considered positive. Additionally, as outlined in methods, we analyzed the complete dataset with a cutoff value of 15 U/mL. Remaining candidate variables were defined similarly to the development dataset.

**Handling of missing values**

We excluded vaccinations with missing values for the following variables: medication data, eGFR, lymphocyte count, hemoglobin.

For the remaining variables, we imputed the median value of the development dataset, since this is the way the online risk calculator would handle missing values as well. Reasons for exclusion and outcome frequency are summarized in **Figure S2.**

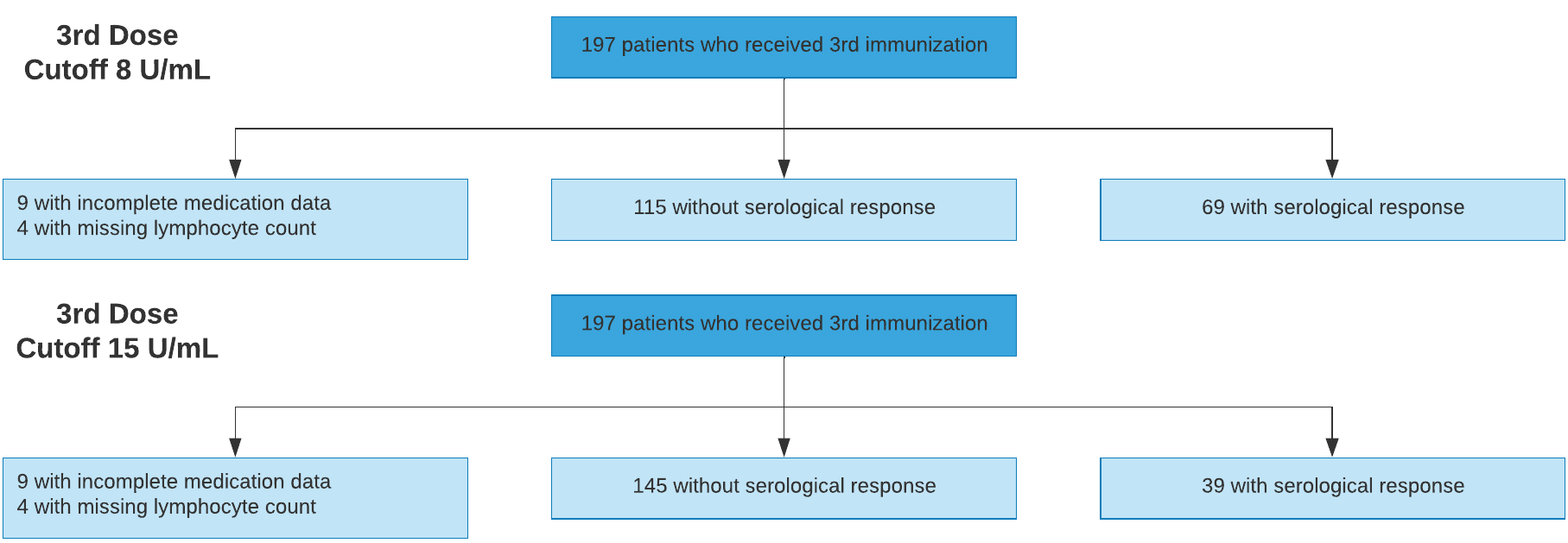

**Figure S2**. Patient flow diagram of the validation cohort 2.

**Item S3.** Detailed information about validation cohort 3

The third validation dataset consisted of 376 vaccinations from the kidney transplant center at the University Hospital of Strasbourg (France), which were derived from a prospective observational trial. All patients provided written informed consent and institutional review board approval was given (approval number: CE-2021-9 and registration number: NCT04828460) for the respective study.

**Outcome and Predictors**

The SARS-CoV-2 assay used was a chemiluminescent microparticle immunoassay (CMIA) SARS-CoV-2 IgG II Quant (Abbott, Rungis, France) with AU/mL as a readout. According to the manufacturer, a value above 50 AU/mL was considered positive. For standardization, AU/mL were converted to BAU/mL according to the manufacturer, with a conversion factor of 1 BAU/mL = 0.142*AU/mL.

Since for most patients, UPCR was available, but not UACR, estimated UACR (eUACR) was calculated from UPCR using the following adjusted equation from Sumida et al.:

eUACR = exp(0.2445 × log(min (UPCR/50, 1)) + 1.5531 × log(max(min(UPCR/500, 1) , 0.1)) + 1.1057 × log(max(UPCR/500, 1)) + 5.2562 – 0.0793 × (if female) + 0.0802 × (if diabetic) + 0.1339 × (if hypertensive)). All patients were assumed to be hypertensive.

**Handling of missing values**

No vaccinations were excluded from analysis, since data on serological response, medication, eGFR, lymphocyte count, hemoglobin contained no missing values. For the remaining variables, we imputed the median value of the development dataset, as described above. Reasons for exclusion and outcome frequency are summarized in **Figure S3**.

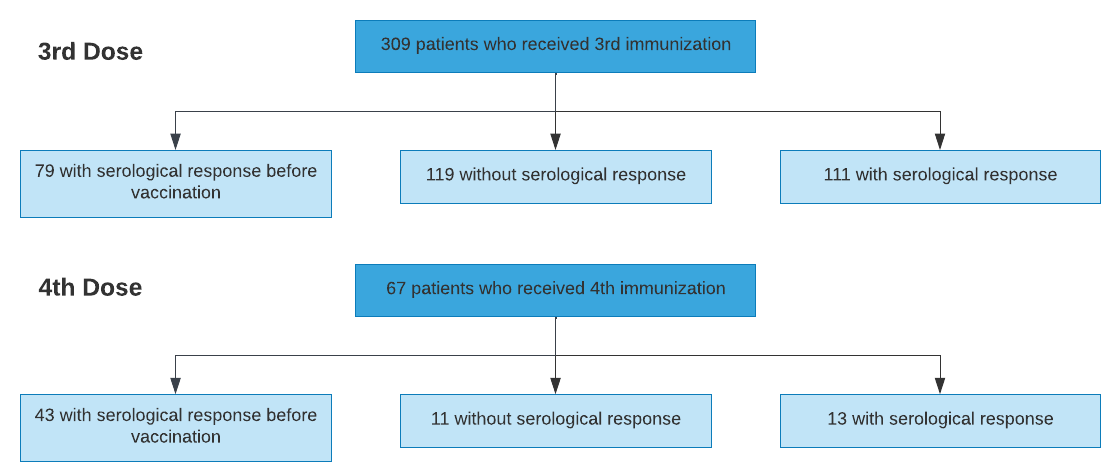

**Figure S3.** Patient flow diagram of the validation cohort 3.

**Item S4.** Detailed information about validation cohort 4

The fourth validation dataset consisted of 904 vaccinations from the kidney transplant center at the Centre Hospitalier Universitaire de Nantes (France). Following informed consent, all patients’ data were extracted from the DIVAT database (Données Informatisées et VAlidées en Transplantation; www.divat.fr, approved by the Comité National de l’Informatique et des Libertés CNIL, No. 914184) and de-identified in order to respect confidentiality. After applying all exclusion criteria, 254 vaccinations were included into the final analysis (cf. Figure S4) when using a cutoff of 0.8 U/mL for electrochemiluminescence immunoassay (ECLIA) (Elecsys, Anti-SARS-CoV-2, Roche Diagnostics GmbH, Mannheim, Germany) and 323 vaccinations were included when using a cutoff of 15 U/mL. The difference is due to the increasing number of vaccinations with pre-vaccination antibody level below the respective positivity cutoff for 15 U/mL.

**Outcome and Predictors**

The SARS-CoV-2 assay used was mostly (206 pre-vaccination measurements and 204 post-vaccination measurements) an electrochemiluminescence immunoassay (ECLIA) (Elecsys, Anti-SARS-CoV-2, Roche Diagnostics GmbH, Mannheim, Germany) with U/mL as a readout. According to the manufacturer, values ≥ 0.8 U/mL were considered positive, and values between the limit of detection (0.4 U/mL) and the positivity cutoff . For secondary analysis, values ≥ 15 U/mL were considered positive, and non-zero values below the cutoff were considered low positive. In 14 pre-vaccination and 20 post-vaccination measurements, the LIAISON® SARS-CoV-2 TrimericS IgG assay (Diasorin, Saluggia, Italy) with a cutoff of ≥ 33.8 BAU/mL was used. In 11 pre-vaccination and 15 post-vaccination measurements, a chemiluminescent microparticle immunoassay (CMIA) SARS-CoV-2 IgG II Quant (Abbott, Rungis, France) with AU/mL as a readout was used, with values ≥ 50 AU/mL considered positive. In 13 pre-vaccination and 12 post-vaccination measurements the NovaLisa SARS-CoV-2 IgG (Novatec Immundiagnostica GmbH, Dietzenbach, Germany), with AU/mL as a readout was used, with values ≥ 11 AU/mL considered positive. In 10 pre-vaccination and 3 post-vaccination measurements, the Atellica® IM SARS-CoV-2 IgG (sCOVG) (Siemens Healthineers, Erlangen, Germany), with cutoff index as a readout was used, with values ≥ 2 considered positive.

Since for most patients, UPCR was available, but not UACR, estimated UACR (eUACR) was calculated from UPCR using the following adjusted equation from Sumida et al.:

eUACR = exp(0.2445 × log(min (UPCR/50, 1)) + 1.5531 × log(max(min(UPCR/500, 1) , 0.1)) + 1.1057 × log(max(UPCR/500, 1)) + 5.2562 – 0.0793 × (if female) + 0.0802 × (if diabetic) + 0.1339 × (if hypertensive)). All patients were assumed to be hypertensive.

**Handling of missing values**

We excluded vaccinations from analysis, for which data on serological response, medication, eGFR, lymphocyte count, or hemoglobin were missing. For the remaining variables, we imputed the median value of the development dataset, since this is the way the online risk calculator would handle missing values as well. Reasons for exclusion and outcome frequency are summarized in **Figure S4**.

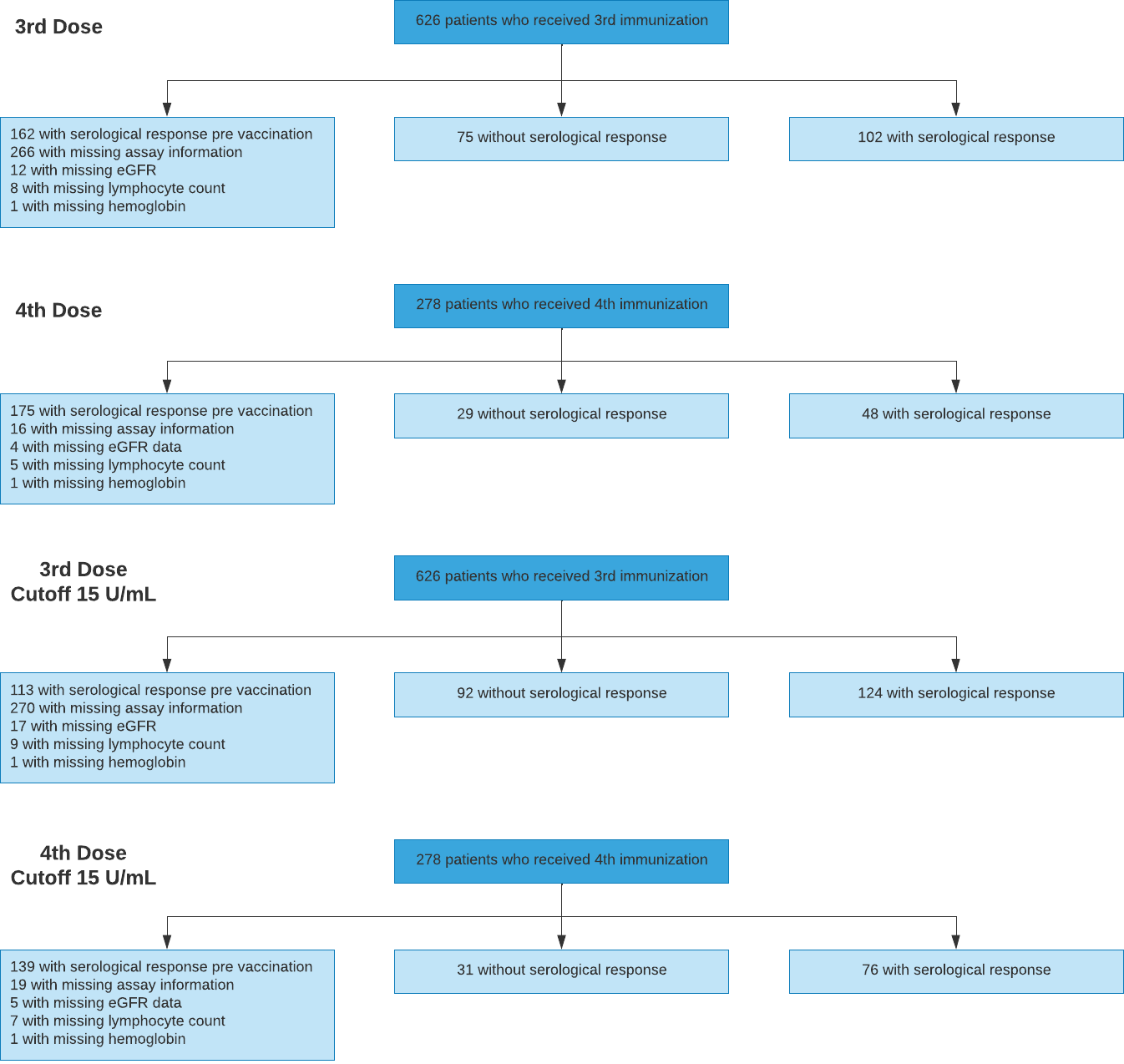

**Figure S4.** Patient flow diagram of the validation cohort 4.

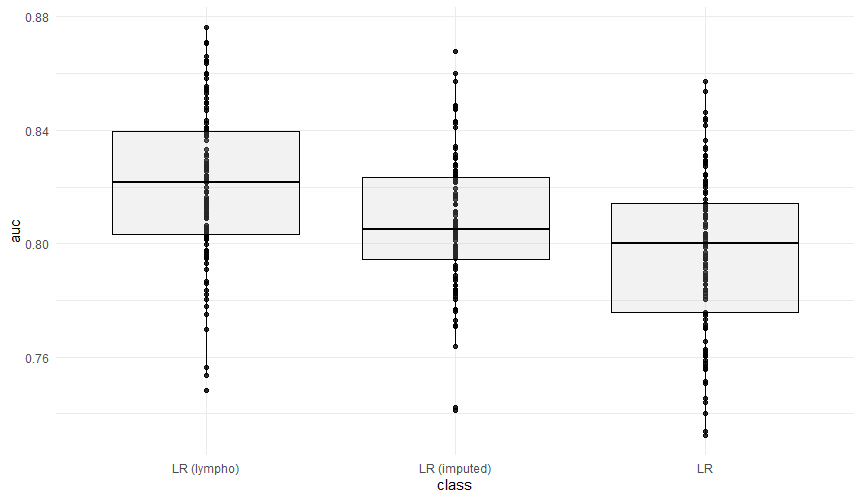

**Figure S5.** Imputation or including patients without lymphocyte count did not achieve better performance during internal validation than complete case analysis. LR (lympho) – logistic regression including lymphocyte count as predictor variable. LR (imputed) – logistic regression with imputed dataset employing multiple imputation. LR – logistic regression without imputation without lymphocyte count as a predictor variable.

**Table S2.** Preliminary analysis did not show improvement of predictive accuracy when excluding lymphocyte count as a predictor variable or by performing multiple imputation for missing laboratory values. Hence, missing lymphocyte count was added as an exclusion criterion. Metrics for predictive accuracy are median AUC-ROC (95% CI), as well as sensitivity (Sens), specificity (Spec), accuracy (Acc.), positive predictive value (PPV), based on the best threshold during ROC-analysis.

| Model Type | AUC | Sens. | Spec. | Acc. | PPV | NPV |
| --- | --- | --- | --- | --- | --- | --- |
| Logistic Regression  with final dataset | 0.822 (0.763 - 0.868) | 0.763 (0.690 - 0.838) | 0.784 (0.679 - 0.861) | 0.778  (0.707 - 0.827) | 0.671 (0.563 - 0.764) | 0.851 (0.795 - 0.898) |
| Logistic regression on dataset without lymphocyte count | 0.800 (0.742 - 0.845) | 0.742 (0.636 - 0.831) | 0.737 (0.652 - 0.837) | 0.742  (0.688 - 0.789) | 0.633 (0.515 - 0.713) | 0.826 (0.769 - 0.879) |
| Logistic regression on dataset using multiple imputation | 0.805 (0.767 - 0.853) | 0.732 (0.649 - 0.832) | 0.756 (0.681 - 0.847) | 0.746  (0.693 - 0.797) | 0.641 (0.557 - 0.729) | 0.826 (0.780 - 0.887) |

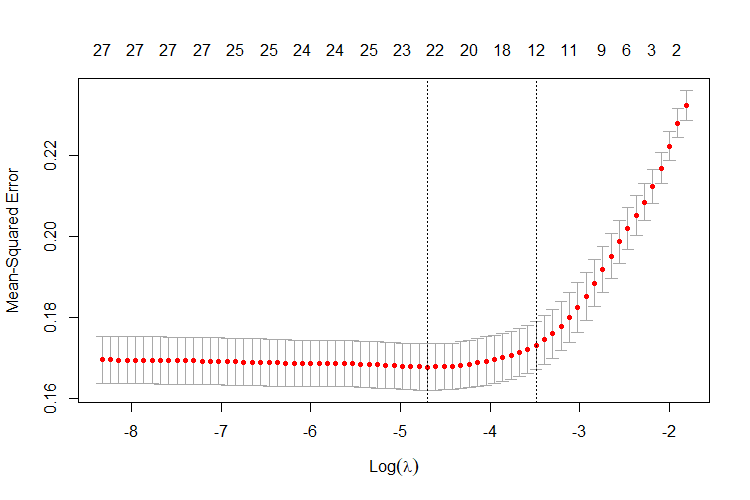

**Figure S6.** Relationship between model sparsity and predictive performance for LASSO logistic regression in the development dataset. For the complete development dataset, the mean squared error (MSE) assessed in an inner five-fold cross-validation is shown in relationship to the hyperparameter λ (bottom) and the number of variables included in the respective model (top). The left dotted line indicates the λ value for which the smallest average MSE is obtained, whereas the right dotted line indicates the largest value of λ for which the MSE remains within one standard error of the minimal MSE. The former one was chosen for the LASSO-Min model and the latter one for the sparser, more regularized LASSO-1SE model.

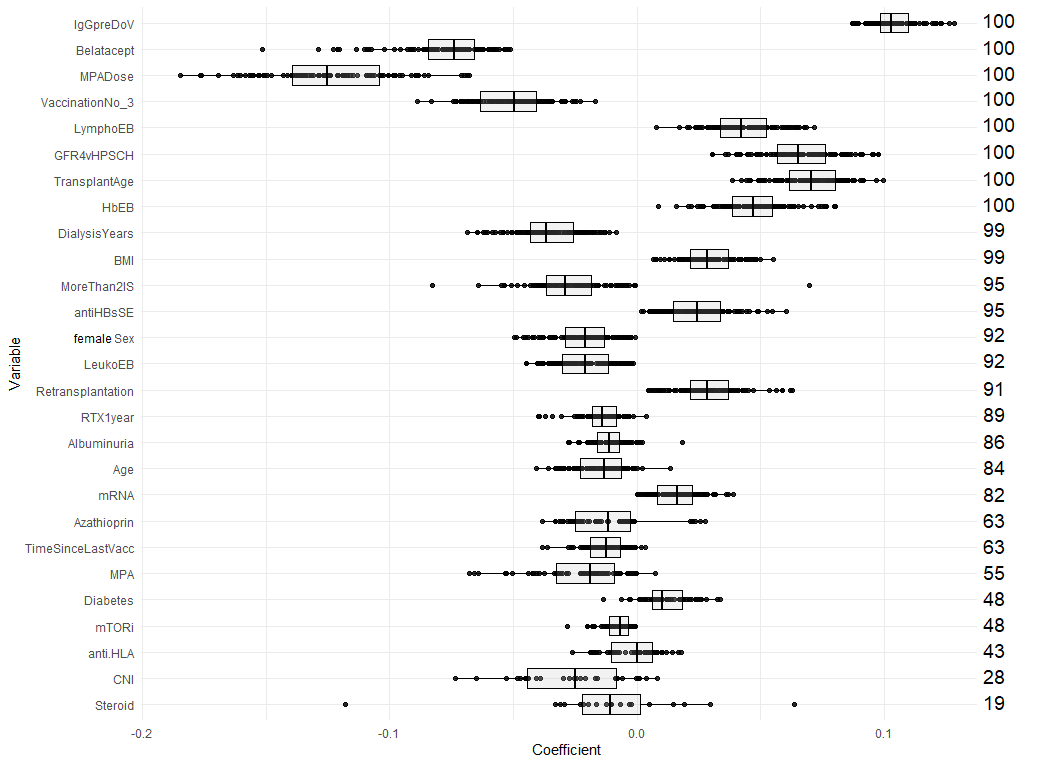

**Figure S7**. Estimated coefficients of the LASSO-Min models summarized across 100 subsampling runs for unstandardized variables. Numbers on the right indicate the selection frequency (in percent) for the respective variable in 100 subsampling runs. Variables are ordered from top to bottom according to the selection frequency.

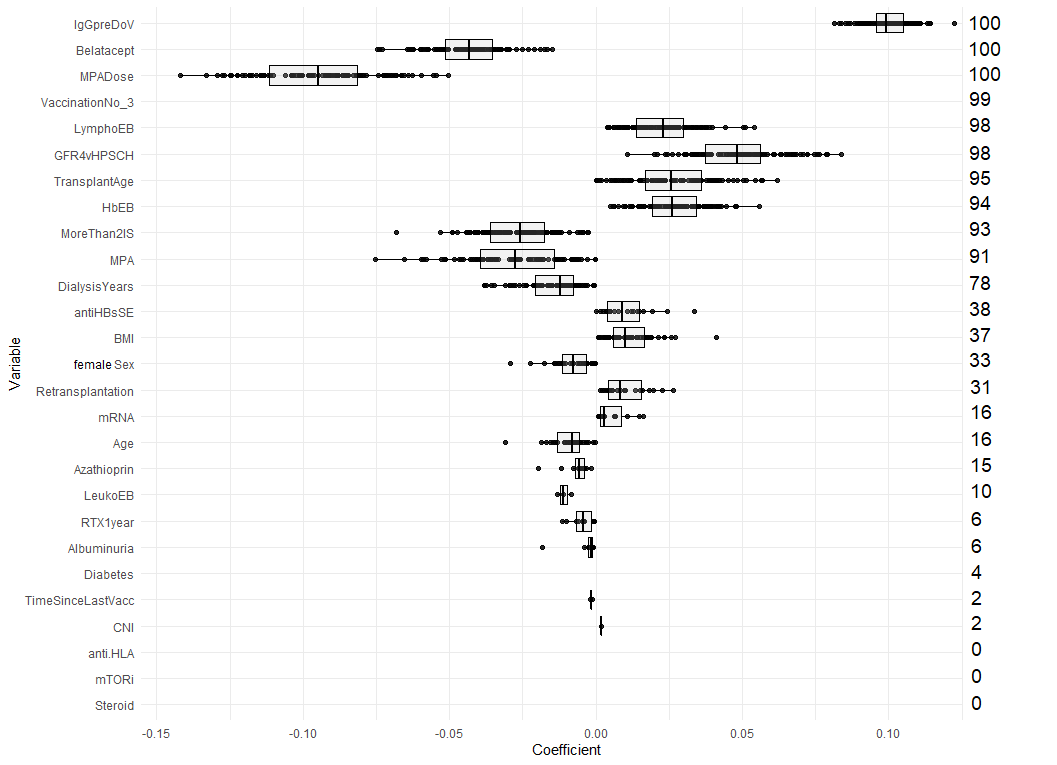

**Figure S8.** Estimated coefficients of the LASSO-1SE models summarized across 100 subsampling runs for unstandardized variables. Numbers on the right indicate the selection frequency (in percent) for the respective variable in 100 subsampling runs. Variables are ordered from top to bottom according to the selection frequency.

**Item S5.** Risk equation for the 11-variable model.

*P_11-var_ (x) = 1 / ( 1 + exp ( - f_11-var_ (x) )*

*f_11-var_ (x) = 0.188049601 + 0.352724365*(Baseline SARS-CoV-2 IgG low positive [0/1]) - 0.085350859*(Third vaccination [0/1]) + 0.004443167*(Transplant age [years]) - 0.002761632*(Dialysis time [years]) - 0.149986747*(Belatacept treatment [0/1]) - 0.070823712*(Mycophenolic acid treatment [0/1]) - 0.105965120*(MPA dose [g MMF equivalent]) - 0.045726834*(More than 2 immunosuppressants [0/1]) + 0.002274646*(Baseline eGFR [ml/min/1.73m2]) + 0.039311344*(lymphocyte count [/nL]) + 0.018128558*(hemoglobin [g/dL])*

**Item S6.** Risk equation for the 23-variable model.

*P_23-var_ (x) = 1 / ( 1 + exp ( - f_23-var_ (x) )*

*f_23-var_(x) = -0.1079327 + 0.4155510*(Baseline SARS-CoV-2 IgG low positive [0/1]) - 0.1124036*(Third vaccination [0/1]) - 0.04148614*(Female dex [0/1]) - 0.001085888*(Age [years]) + 0.007465119*(BMI [kg/m2]) + 0.02857272*(mRNA vaccination [0/1]) + 0.1343634*(Retransplantation [0/1]) + 0.01113518*(Transplant age [years]) - 0.007937533*(Dialysis time [years]) - 0.2339575*(Belatacept treatment [0/1]) - 0.008675321*(Mycophenolic acid treatment [0/1]) - 0.1773556*(MPA dose [g MMF equivalent]) - 0.007156921*(mTOR inhibitor treatment [0/1]) - 0.03860331*(Azathioprine treatment [0/1]) - 0.045726834*(More than 2 immunosuppressants [0/1]) - 0.01829209*(Rituximab in the last year [0/1]) - 0.0002238902*(Time since previous vaccination [days]) + 0.003344195*(Baseline eGFR [ml/min/1.73m2]) - 0.007810127*(leukocyte count [/nL]) + 0.06496441*(lymphocyte count [/nL]) + 0.02987510*(hemoglobin [g/dL]) + 0.00006856821*(Anti-HBs [U/mL]) - 0.01705246*(albumin-creatinine ratio [g/g])*

**Table S3.** Performance of the 23-variable model during external validation. AUC-ROC, as well as sensitivity (Sens), specificity (Spec), accuracy (Acc.), positive predictive value (PPV), negative predictive value assessed on each validation set. The threshold was derived during ROC-analysis on the development dataset. To provide 95% CI, empirical 2.5% and 97.5% quantiles of the respective metric are provided after performing a 1000-fold nonparametric ordinary bootstrapping with each validation set.

| Model Type | AUC  point estimate (95% CI) | Sens  point estimate (95% CI) | Spec  point estimate (95% CI) | Acc  point estimate (95% CI) | PPV  point estimate (95% CI) | NPV  point estimate (95% CI) |
| --- | --- | --- | --- | --- | --- | --- |
| Validation 1  23-variable | 0.853 (0.795 - 0.908) | 0.815  (0.718 - 0.906) | 0.643  (0.555 - 0.727) | 0.702  (0.634 - 0.764) | 0.541  (0.450 - 0.644) | 0.871  (0.800 - 0.932) |
| Validation 2  23-variable  (cutoff 0.8/mL) | 0.692  (0.613 - 0.763) | 0.290  (0.179 - 0.391) | 0.826  (0.750 - 0.889) | 0.625  (0.549 - 0.690) | 0.500  (0.342 - 0.651) | 0.660  (0.577 - 0.735) |
| Validation 2  23-variable  (cutoff 15 U/mL) | 0.714  (0.627 - 0.787) | 0.385  (0.237 - 0.531) | 0.828  (0.769 - 0.883) | 0.734  (0.668 - 0.793) | 0.375  (0.231 - 0.528) | 0.833  (0.767 - 0.890) |
| Validation 3  23-variable | 0.844  (0.789 - 0.889) | 0.823  (0.750 - 0.891) | 0.677  (0.595 - 0.756) | 0.748  (0.689 - 0.799) | 0.708  (0.635 - 0.783) | 0.800  (0.720 - 0.871) |
| Validation 4  23-variable  (cutoff 0.8/mL) | 0.730 (0.668 - 0.790) | 0.773 (0.711 - 0.842) | 0.606 (0.505 - 0.699) | 0.705 (0.650 - 0.756) | 0.739 (0.669 - 0.807) | 0.649 (0.563 - 0.740) |
| Validation 4  23-variable  (cutoff 15 U/mL) | 0.795 (0.741 - 0.843) | 0.855 (0.804 - 0.900) | 0.544 (0.459 - 0.633) | 0.737 (0.690 - 0.780) | 0.753 (0.696 - 0.805) | 0.698 (0.594 - 0.787) |
| **Overall performance**  **23-variable** | **0.776 (0.744 - 0.807)** | **0.713 (0.666 - 0.760)** | **0.688 (0.646 - 0.730)** | **0.700 (0.668 - 0.729)** | **0.663 (0.617 - 0.708)** | **0.736 (0.693 - 0.778)** |
| **Overall performance**  **23-variable**  **(cutoff 15 U/mL)** | **0.827 (0.801 - 0.854)** | **0.797 (0.757 - 0.835)** | **0.679 (0.642 - 0.720)** | **0.732 (0.704 - 0.761)** | **0.670 (0.630 - 0.712)** | **0.804 (0.767 - 0.842)** |

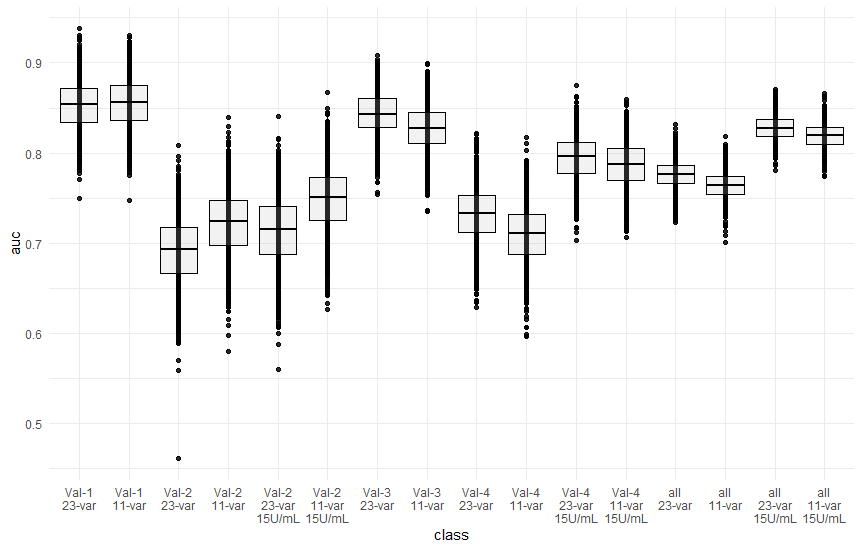

**Figure S9.** Predictive performance of both the 23-variable and 11-variable models in four independent external validation cohorts.
